# A Transcriptomic Roadmap of Parkinson’s Disease Progression at Single Cell Resolution

**DOI:** 10.1101/2025.07.30.25332436

**Authors:** Tereza Clarence, Nicolas Masse, Christian Porras, NM Prashant, Sabina Berretta, Vahram Haroutunian, David A. Davis, William K. Scott, Gabriel Hoffman, John Fullard, Donghoon Lee, Jaroslav Bendl, Panos Roussos

## Abstract

Parkinson’s disease (PD) is a progressive neurodegenerative disorder with complex and heterogeneous molecular pathology across the brain. However, the full cellular architecture of PD progression remains unresolved. Here, we present a comprehensive single-nucleus transcriptomic atlas of PD spanning multiple anatomically and clinically relevant brain regions to capture shared and region-influenced transcriptomic features from 97 deeply phenotyped donors, including individuals across the full spectrum of Braak Lewy body stages. Profiling over 2 million nuclei, we define 62 transcriptionally distinct cell subtypes and uncover widespread, cell-type-specific gene expression changes across neurons, glia, and vascular cells. We identify convergent upregulation of stress-responsive transcriptional programs - such as unfolded protein response, DNA damage repair, and autophagy - across multiple cell types, with key regulators including HSF1, MYC, and FOXO3. Integrative analyses link these transcriptional alterations to PD genetic risk, revealing transcription factor to target gene networks enriched for PD GWAS loci in microglia and neuronal subpopulations. To quantify disease burden at cellular resolution, we introduce a transcriptomic pathology score, revealing early-stage activation in neurons and myeloid cells, followed by delayed engagement of vascular populations. We further demonstrate that microglia undergo dynamic, subtype-specific transitions across disease stages, including early adaptive responses and late-phase stress and proliferative programs. Altered cell-cell communication networks, particularly involving myeloid–neuronal signaling, highlight a progressive rewiring of neuroimmune interactions. This atlas provides a foundational resource for understanding PD progression at single-cell resolution, linking genetic risk to dynamic molecular pathology, and illuminating stage-specific targets for therapeutic intervention.

## Introduction

Parkinson’s disease (PD) is a progressive neurodegenerative disorder characterized primarily by motor symptoms, such as tremors, rigidity, and bradykinesia, which arise from the loss of dopaminergic neurons in the substantia nigra (SN) of the midbrain^1^. While the substantia nigra has been the primary focus of PD research, it is increasingly recognized that PD is a multifaceted disorder involving widespread brain regions and complex molecular mechanisms that extend far beyond the SN. PD progresses through the brain in a pattern described by Braak Lewy body (LB) staging^2,3^, in which the spread of Lewy pathology closely mirrors disease severity and clinical symptomatology. However, the full molecular and transcriptomic response to this disease propagation remains incompletely characterized, leaving critical gaps in our understanding of PD progression.

PD is increasingly recognized as a complex disorder that extends beyond dopaminergic neuron degeneration in SN. Histopathological and transcriptomic studies have revealed that multiple brain cell types, including astrocytes, microglia, oligodendrocytes, and vascular cells, exhibit disease-related alterations, implicating a broad spectrum of cell types in PD pathogenesis^4,5^. For instance, microglial activation^6^, astrocyte dysfunction^7,8^, and endothelial barrier breakdown have all been reported in PD and are thought to contribute to both neuroinflammation and neuronal vulnerability^9^. These findings emphasize the need to systematically resolve how PD affects diverse cellular compartments across brain regions. Single-cell and single-nucleus RNA sequencing (scRNA-seq/snRNA-seq) technologies now provide a powerful means to dissect these complex, cell-type-specific transcriptional changes at scale. While these approaches have yielded valuable insights, particularly into dopaminergic neuron vulnerability in the SN^10,11^, they have been less frequently applied to extra-nigral regions. Yet, many non-motor symptoms of PD, such as cognitive decline, mood disorders, and autonomic dysfunction, are rooted in cortical^12,13^ and brainstem dysfunction^14^. Recent work has also emphasized the importance of glial and immune responses in these regions, showing that glial cells can modulate neuronal function and suppress ferroptosis-mediated neurodegeneration^15^. Thus, there remains a critical need for multi-regional, cell-type-resolved studies to elucidate the full cellular architecture of PD pathology.

To address this gap, we performed in-depth analyses of a multi-brain region, single-nucleus transcriptomic dataset, previously generated by our lab^16^, comprising a cohort of 444 samples from 97 donors, spanning various stages of PD progression as reflected by regional spread of Lewy bodies throughout the brain. This expansive dataset enabled the systematic investigation of transcriptomic responses, shifts in cellular abundance, and altered cell-cell interactions throughout PD progression. In addition, we investigated the dynamic behavior of microglia, the brain’s resident immune cells, across stages of PD, focusing on their transcriptional and compositional changes in the context of neuroinflammation and neurodegeneration^8,17,18^. By capturing these spatial and cellular dynamics, our analyses provide deeper biological insights into PD and serve as a foundation for future studies aimed at uncovering disease mechanisms and developing more precise therapeutic strategies.

## Results

### Multi-regional single-nucleus transcriptomic atlas of Parkinson’s disease

We generated a multi-regional single-nucleus transcriptomic atlas of PD using postmortem brain tissue from 97 donors enrolled in the Accelerating Medicines Partnership for Parkinson’s Disease (AMP-PD) consortium (**Fig. 1a**, **Supplementary** Fig. 1a-b, **Supplementary Table 1**), including preclinical, clinically defined PD and control cases, capturing a broad spectrum of Lewy body (LB) pathology, as measured by Braak LB staging^2,3^ (range 0-6). The cohort spans late adulthood (ages 57-102 years) and includes a balanced distribution of male and female donors, and diverse ancestry groups (**Extended Data** Fig. 1a-b, **Supplementary** Fig. 1a-c).

**Figure 1.**
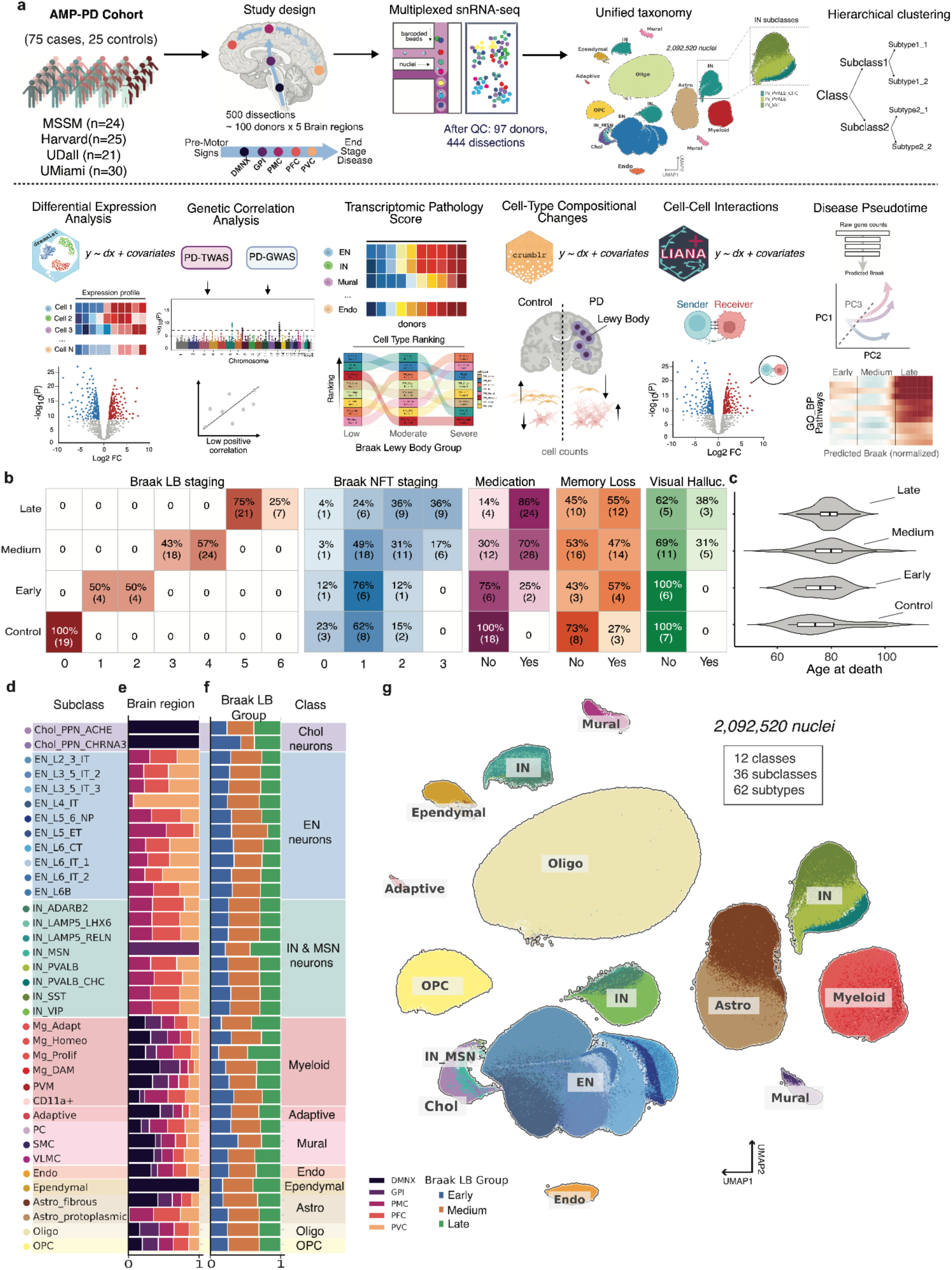
Multi-regional single-nucleus transcriptomic atlas of Parkinson’s disease progression. **(a)** *Study design;* The upper panel illustrates the cohort structure and experimental workflow^16^, including multi-regional snRNA-seq and construction of a hierarchical taxonomy with three levels of cellular granularity: class, subclass, and subtype. The lower panel (below the dashed line) summarizes downstream analyses performed on the multi-regional atlas, including: differential expression (DE) analysis using dreamlet^23^, genetic correlation of DE signatures with PD TWAS and GWAS results, transcriptomic pathology scoring (TPS) of individual donors, assessment of cell type composition changes using crumblr^24^, inference of cell-cell interactions via the LIANA+ pipeline^25^, and exploration of disease progression through pseudotime analysis. **(b-c)** *Clinical metadata overview;* Panel (b) visualizes available clinical and neuropathological metadata, including Braak Lewy Body (LB) stage as a measure of PD pathology (shades of red), Braak neurofibrillary tangle (NFT) stage as a measure of Alzheimer’s pathology (shades of blue), PD medication (shades of magenta), memory loss (shades of orange), and presence of visual hallucinations (shades of green). Panel (c) shows the distribution of age across Braak LB groups, categorized as Control, Early (Braak LB 1-2), Medium (Braak LB 3-4), and Late (Braak LB 5-6). Percentages are based only on donors for whom clinical data were available. **(d-f)** Stacked bar plot showing the contribution of different brain regions (e) and Braak LB group (f) to each subclass (d). **(g)** UMAP embedding visualization of the AMP-PD dataset, colored by subclass identity, consistent with the taxonomy shown in panel (d-e).

We focused on five brain regions, affected progressively according to Braak LB staging chronology, including the dorsal motor nucleus of the Xth nerve (DMNX), globus pallidus interna (GPI), primary motor cortex (PMC), prefrontal cortex (PFC), and primary visual cortex (PVC). These regions collectively represent critical nodes in the neural circuitry implicated in PD, and their study is essential for a comprehensive understanding of the disease’s pathophysiology. In addition to PD neuropathological annotations, further clinical metadata were available, including Braak neurofibrillary tangle (NFT) staging^19^ as a measure of co-occurring Alzheimer’s disease (AD) pathology, as well as clinical indicators such as PD medication usage, presence of visual hallucinations, and memory loss (**Fig. 1b**, **Supplementary Table 1**). SnRNA-seq was performed across the DMNX, GPI, PMC, PFC, and PVC (**Extended Data** Fig. 1c-d).

To ensure cross-donor and cross-region comparability, we implemented a unified experimental and computational pipeline (**Methods**). After rigorous quality control, 2,092,520 nuclei and harmonized clinical and technical metadata were retained for downstream analysis.

We generated a hierarchical taxonomy of brain cell types by integrating both joint (cross-regional) and region-specific clustering, allowing us to capture anatomical diversity while maintaining a unified classification framework (**Extended Data** Fig. 1e). The hierarchical taxonomy spans three levels of granularity - class, subclass, and subtype - comprising 12 major classes: Excitatory Neurons (EN), Inhibitory Neurons (IN), Cholinergic Neurons (Chol), Medium Spiny Neurons (MSN), Myeloid cells, Mural cells, Adaptive Immune cells, Endothelial cells (Endo), Ependymal cells, Astrocytes (Astro), Oligodendrocytes (Oligo), and Oligodendrocyte Precursor Cells (OPCs). These cell type classes are further divided into 36 subclasses and 62 subtypes (**Fig. 1d-f**). Taxonomy annotations were aligned and validated against previously published datasets from the PsychAD cohort^20^ and the Alzheimer’s disease (AD) dataset from Mathys et al. (2023)^21^ (**Extended Data** Fig. 1Ff**-g**, **Supplementary** Fig. 3**, Supplementary Table 2**), and demonstrated robust expression of canonical marker genes (**Supplementary** Fig. 2).

Region-specific mapping across the class, subclass, and subtype levels of the taxonomy revealed substantial neuronal heterogeneity across brain regions (**Fig. 1d-e, Supplementary** Fig. 4). Notably, Chol neurons were found exclusively in the DMNX (**Extended Data** Fig. 2a-b), while MSN neurons were restricted to the GPI (**Extended Data** Fig. 2a, c). In contrast, cortical regions (PMC, PFC, and PVC) shared the majority of EN and IN neuronal subclasses (**Extended Data** Fig. 2d-f). Glial and vascular cell types were largely conserved across all regions, with the exception of ependymal cells, which were enriched predominantly in the DMNX.

We systematically examined disease-specific associations at the subclass level of our cellular taxonomy using scDRS tool^22^ across selected neurological, metabolic, immune and psychiatric traits (**Extended Data** Fig. 3, **Methods, Supplementary Table 3**). Notably, PD exhibited strong and selective enrichment within multiple Micro subclasses, including Mg_DAM, Mg_Adapt, Mg_Homeo, and Mg_Prolif, implicating innate immune cells as key mediators of genetically driven transcriptomic dysregulation in PD. In contrast, neuronal subclasses demonstrated more trait-specific association profiles, particularly across psychiatric and cognitive traits. Subclasses of IN and EN exhibited distinct enrichment patterns for disorders such as schizophrenia, major depressive disorder, and educational attainment, highlighting a broader organization of genetic risk that is closely aligned with neuronal identity and function across neuropsychiatric and neurodevelopmental disorders.

This high-resolution, multi-regional single-nucleus transcriptomic dataset provided a comprehensive framework for systematically investigating cell type-specific molecular vulnerability in PD. In the following sections, we leverage this resource to explore differential gene expression, compositional dynamics, transcriptional regulatory programs, genetic convergence with PD risk loci, alterations in the intercellular communication landscape, and trajectories of disease progression across diverse brain cell types.

### Cell Type-Specific Dysregulation of Biological Pathways and Transcriptional Networks in PD

We identified differentially expressed genes (DEGs) between PD and control subjects within each cell type using a linear mixed model framework implemented in dreamlet^23^, after adjusting for technical and biological covariates (**Methods, Supplementary Table 4**). The effect of biological and technical covariates that contribute to gene expression variability is estimated based on variance partition^28^ analysis (**Supplementary** Fig. 5a-c). We identified 11,276 unique DEGs among 26,721 total DEGs (FDR < 0.05) across all cell types between PD cases and controls. Of these, 7,219 DEGs (64% of unique DEGs) were dysregulated across multiple cell types, while 4,057 DEGs (36% out unique DEGs) were cell-type specific (**Supplementary** Fig. 5d-e), reflecting both shared and cell-type–specific transcriptional alterations in PD.

We observed strong concordance between differential expression results from PD vs. Control and Braak LB staging analyses (**Supplementary** Fig. 6a), indicating consistent disease-associated transcriptional signatures across comparisons; for subsequent analyses, we focused on PD vs. Control results, which exhibited greater log_2_FC magnitudes and clearer case-control contrasts. The distribution of DEGs across cell types revealed a relatively even balance between up- and down-regulated genes (**Supplementary** Fig. 6b). The largest number of significant DEGs (FDR < 0.05, |log_2_FC| > 0.2) was observed in ENs and Astros, followed by Oligos. While cluster size may contribute to DEG counts to some extent, the observed transcriptional responses likely reflect cell-type-specific vulnerabilities in PD (**Supplementary** Fig. 6c). Notably, HSPA1A and its related heat shock protein (HSP) family genes are found to be upregulated in Mural, Endo, and OPC cells (**Fig. 2a**), suggesting activation of protective proteostasis mechanisms. These molecular chaperones help maintain protein homeostasis by promoting correct folding and clearance of misfolded proteins. In PD, α-synuclein aggregation disrupts this balance, overwhelming the chaperone system and contributing to neuronal vulnerability and degeneration^29^.

**Figure 2.**
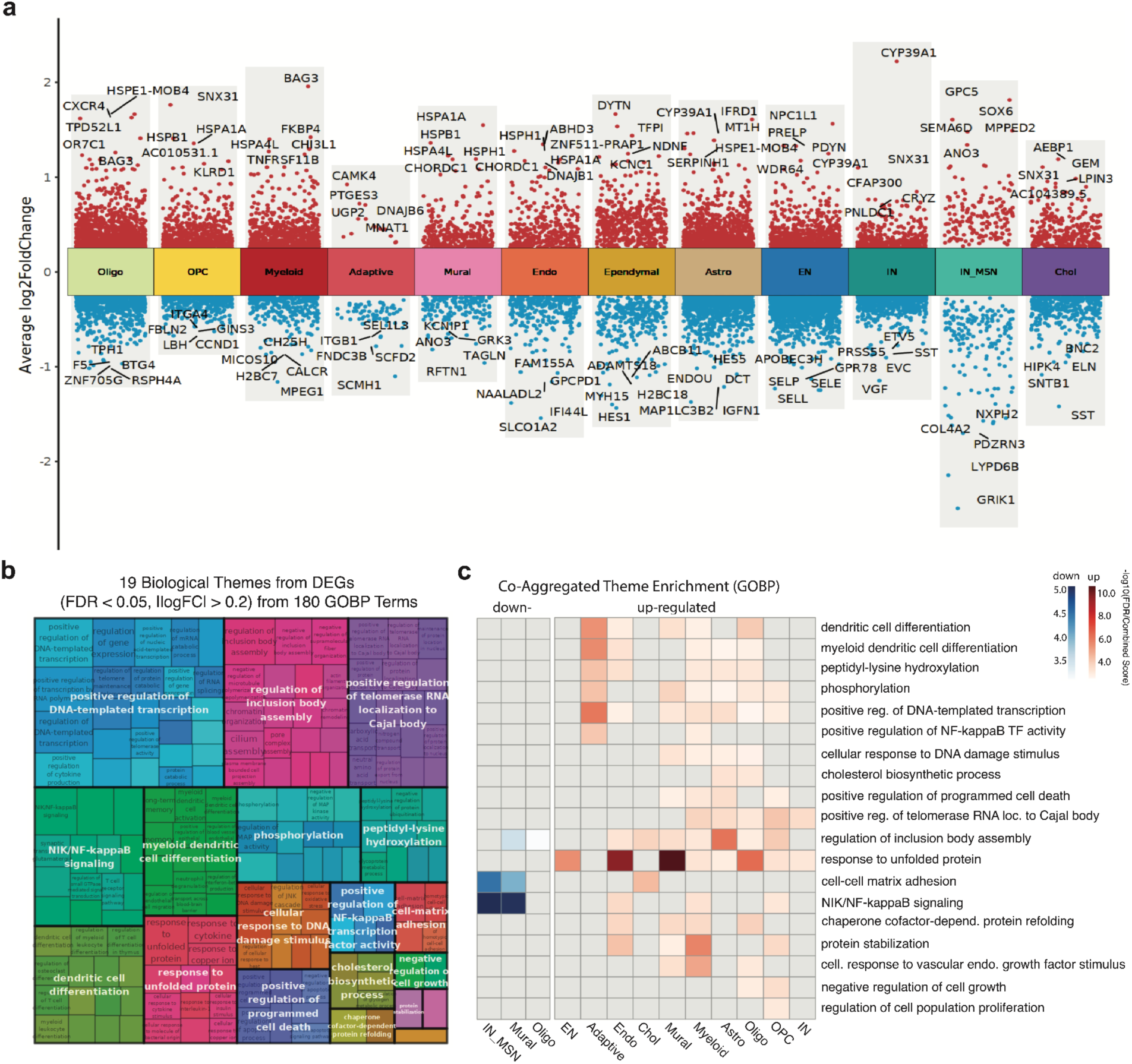
Cell type-specific gene expression changes in PD. (a) *Differentially expressed genes (DEGs) per cell class*, illustrating the range of log_2_FC. Red indicates up-regulated DEGs, while blue represents down-regulated DEGs. Genes with the highest absolute log_2_FC values are labeled. (b) *Enrichment themes of biological processes* (FDR < 0.05) derived from cell type-specific up- and down-regulated DEG sets via gene set enrichment analysis (GSEA). Gene ontology (GO) terms are clustered into biological themes, with small boxes representing individual GO terms and large colored boxes corresponding to broader functional categories, named after their most significantly enriched GO term (**Supplementary Table 5**). (c) *Aggregated enrichment of GO terms within biological themes*, stratified by cell type-specific up-regulated (red) and down-regulated (blue) DEGs. Cell types without significant enrichment for key biological themes are not listed.

To evaluate the functional consequences of PD-associated gene dysregulation in specific cell types, we performed Gene Set Enrichment Analysis (GSEA) for DEGs with |log_2_FC| > 0.2 and FDR < 5% using the GO Biological Process (GOBP) database (**Methods**)^30^ to prioritize biologically meaningful pathways while maintaining analytical rigor (see **Methods**). To facilitate biological interpretation, 180 significant GOBP terms (FDR <0.05) were semantically clustered into 19 biological themes (**Fig. 2b-c**, **Supplementary Table 5, Methods**^31^). The largest biological themes centered around “positive regulation of DNA-templated transcription” (GO:0045893), “regulation of inclusion body assembly” (GO:0090083) and “positive regulation of telomerase RNA localization to the Cajal body (GO:1904874)”^32,33^ (**Fig. 2b**), which encompassed pathways related to amino acid transport and intracellular RNA localization, possibly reflecting dysregulated cellular trafficking mechanisms in PD. Other pathology-related themes included “cellular response to DNA damage stimulus” (GO:0006974), “response to unfolded protein” (GO:0006986) and “positive regulation of programmed cell death” (GO:0043068).

We investigated cell type specificity and directional changes in these biological themes by mapping the FDR-significant GOBP terms within each theme to the specific cell types in which they were enriched (**Fig. 2c**). While the overall numbers of upregulated and downregulated DEGs were relatively balanced across cell types, the majority of enriched biological pathways were upregulated (**Fig. 2c**). Neuronal and non-neuronal populations demonstrated distinct enrichment patterns, with neurons predominantly engaging in unfolded protein responses, whereas myeloid, astro and oligo cells exhibited widespread enrichment across multiple biological themes. Furthermore, processes such as “response to unfolded protein” and “regulation of inclusion body assembly” were dysregulated across the largest number of cell types, suggesting shared vulnerability across distinct cellular populations in PD.

To assess the convergence between PD-associated genetic risk and disease-related transcriptional dysregulation, we then examined the overlap among 84 protein-coding genes prioritized by DEGs (**Extended Data** Fig. 4**, Supplementary Table 6**) and recent PD genome-wide association studies (GWAS)^34^. For each gene, we assigned a putative cell type based on the highest absolute t-statistic across taxonomy classes, a strategy supported by strong concordance between DE comparisons (Spearman ρ = 0.91, p < 0.05 for PD vs. control and Braak LB staging comparisons; **Supplementary** Fig. 7a).

Genes prioritized for Myeloid and Oligo cells were predominantly upregulated, while those mapped to Astros, OPCs, ENs and INs showed a more balanced distribution of up- and downregulated expression. Notably, 18 of the 84 genes exhibited consistent directionality across all cell types in which they were detected, suggesting potential pleiotropic or context-dependent regulation. These included *DDRGK1*, *CRLS1*, *SPPL2B*, *NOD2*, *RPS6KL1*, *CAB39L*, *RNF141*, *GBF1*, *BAG3*, *FGF20*, *HLA-DRB5*, *FAM47E-STBD1*, *BST1*, *TMEM175*, *FAM47E*, *SPTSSB*, *PMVK*, and *FCGR2A*. To further contextualize these findings, we annotated each GWAS gene by functional classification, transcriptional directionality, chromosomal locus, and associated genetic context (**Extended Data** Fig. 4). Functional grouping revealed enrichment in biological categories highly relevant to PD pathophysiology, including vesicle trafficking, RNA processing, synaptic signaling, mitochondrial dynamics, and immune-related pathways (**Supplementary** Fig. 7b-c).

### Integrative TF-target gene network analysis reveals convergence of transcriptional and genetic signals in PD

To gain insight into the regulatory context of the differentially expressed genes, we applied transcription factor (TF) inference (**Fig. 3a**; **Supplementary Table 7, see Methods**), which inferred TF activity scores per cell type and identified top upregulated, cell type-specific TFs in PD (**Fig. 3b**). Several TFs demonstrated strong cell-type-specific enrichment and formed interconnected TF-TF protein-protein interaction (PPI) networks (**Supplementary** Fig. 8a). A subset of TFs, HSF1, FOXO3, MYC, and AR, were consistently upregulated across multiple cell types and exhibited strong connectivity within TF-TF PPI networks (**Supplementary** Fig. 8b-c), suggesting they may constitute a core regulatory module in PD pathogenesis. Interestingly, these TFs have known roles in neurodegeneration, including HSF1 in proteostasis and stress response^35–37^, FOXO3 in oxidative stress resilience^38–41^, MYC in cell cycle regulation^42–44^, and AR in hormonal modulation and neuroinflammation^45–47^.

**Figure 3.**
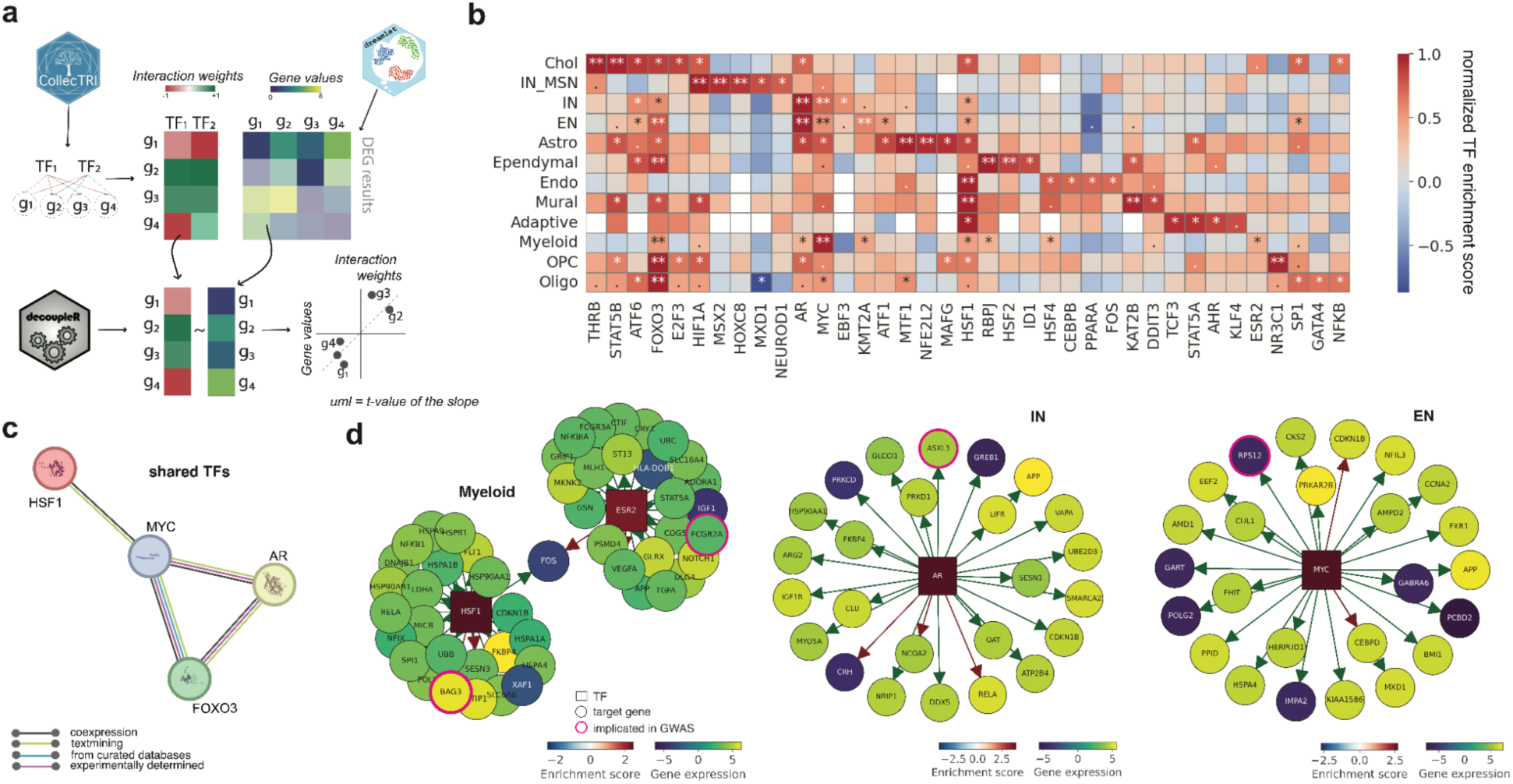
Regulatory network inference and overlap with GWAS genes reveals shared and cell type-specific transcriptional programs in PD. **(a)** *Schematic of transcription factor (TF) inference using decoupleR*^48^, integrating DEGs with a univariate linear model (UML) approach and the CollecTri TF database^49^. The method integrates TF-target interaction weights with gene expression values from differential expression analysis and infers TF activity per cell type using a univariate linear model. Modified from Pau Badia-i-Mompel et al. (2022)^48^ with permission from authors. **(b)** *Top five TFs with the highest cell-type specificity and enrichment scores* for each cell type. Color intensity corresponds to the normalized TF enrichment score, with statistical significance denoted as follows: ** *FDR < 0.01*, * *FDR < 0.05*, and · *FDR < 0.1*. **(c)** *Network of TFs consistently enriched across multiple cell types* (HSF1, MYC, AR, FOXO3). **(d)** *Visualization of selected TF-target gene networks* for ESR2 and HSF1 in Myeloid, AR in IN, and MYC in EN cell types. Node fill color represents gene expression and border halo indicates GWAS-implicated genes. Arrow direction reflects TF → gene regulation, with edge color indicating directionality (positive or negative effect).

Given the potential role of TFs in mediating genetic risk for PD, we sought to determine whether TF-target gene networks are enriched for PD-associated loci. To this end, we systematically intersected PD GWAS-prioritized genes with TF-target gene networks previously defined from upregulated TFs across all cell types (see **Fig. 3b**). In total, 9 GWAS-implicated genes (*BAG3*, *MAP3K14*, *FCGR2A*, *UBTF*, *ASXL3*, *IP6K2*, *RPS12*, *GCH1*, *CAMK2D,* **Supplementary Table 8**) were recovered across TF-target gene networks (**Fig. 3c**, **Supplementary** Fig. 8d). With the exception of *UBTF*, all recovered genes were identified within TF-target gene networks derived from the same cell type that was prioritized by DE analysis, strengthening the cell-type specificity and robustness of this integrative framework. Furthermore, 7 out 9 GWAS genes were uniquely assigned to one TF-target gene cell type network, reinforcing the biological specificity of these associations. The exceptions were *BAG3* and *MAP3K14* genes, found in HSF1 TF-target gene networks across multiple cell types (Myeloid, Astro, OPC, neuronal populations), suggesting a potential pleiotropic effect. Interestingly, the role of HSF1 has been implicated in modulation of microglia activation and has recently gained popularity as a potential therapeutic target for PD^36,37^

Finally, we visualized representative TF-target gene networks from HSF1 and ESR2, AR and MYC TFs in relevant cell types (**Fig. 3d**). These networks illustrate both the expression patterns and regulatory influence of key TFs, providing a cell type-resolved view of transcriptional control over *BAG3*, *FCGR2A*, *ASXL3* and *RPS12* genes implicated in PD genetic risk.

### Transcriptomic Pathology Score recapitulates heterogeneity in PD population

Understanding how transcriptional changes relate to clinical and pathological features of PD at the individual level is critical for identifying molecular markers of disease progression. Traditional case-control comparisons may obscure the heterogeneity of transcriptomic alterations across individuals and cell types. To address this, we utilized the Transcriptomic Pathology Score (TPS)^51^, a continuous metric that quantifies the similarity between an individual’s gene expression profile and a PD-associated differential expression signature. This approach enables refined assessment of transcriptomic dysregulation, moving beyond traditional binary case-control classification. By analyzing TPS across brain regions, cell types, and diverse PD-related clinical, genetic, and neuropathological measures, we aimed to uncover cell-type- and subject-specific molecular correlates of disease severity and progression (**Fig. 4a-b**, **Methods**).

**Figure 4.**
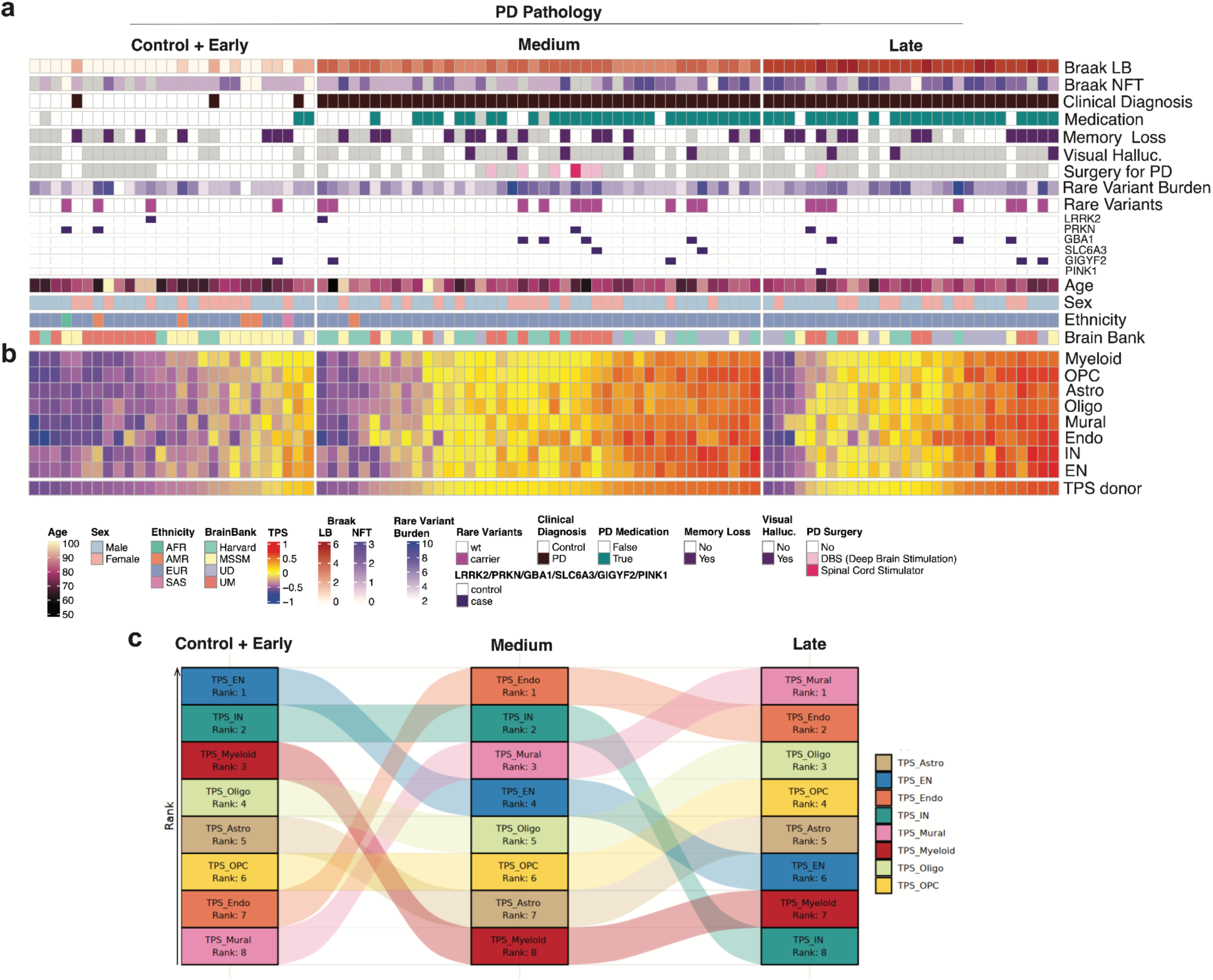
Transcriptomic Pathology Score (TPS) across Braak LB stages at the donor and cell-type level. **(a)** Overview of demographics (age, sex, ethnicity, brain bank), rare variant burden, rare variant carriers for *LRRK2*, *PRKN*, *GBA1*, *SLC6A3*, *GIGYF2*, and *PINK1*), as well as neuropathological metrics of PD progression (Braak Lewy body; Braak LB), AD progression (Braak neurofibrillary tangles; Braak NFT), indication of memory loss, clinical diagnosis (PD vs. control), and PD treatment. **(b)** *Average TPS scores per donor and cell-type specific TPS*, grouped by Early+Control, Medium, and Late Braak LB stages. Each column represents an individual donor, while each row corresponds to a specific cell type. Warmer colors indicate higher TPS values, suggesting an increase in transcriptomic pathology in specific cell types with disease progression. **(c)** *Alluvial plot showing the ranking of cell types based on their mean TPS* within each Braak LB stages (Control+Early, Medium, Late). This visualization highlights the progressive involvement of specific cell types in PD pathology.

TPS values were highly correlated across brain regions (average Spearman ρ = 0.967 and average Pearson *r* = 0.975; **Supplementary** Fig. 9a), suggesting a consistent transcriptional signature associated with PD pathology across distinct brain regions. Additionally, TPS was strongly positively correlated with both Braak LB and Braak LB, further supporting its utility as a transcriptomic marker of PD-related pathological burden. Genetic variation also contributed to differences in TPS, as individuals with a higher burden of rare coding variants exhibited elevated TPS values, which were positively correlated with Braak LB staging (Spearman ρ = 0.309, **Supplementary** Fig. 9a, c). At the cell-type level (**Supplementary** Fig. 9b), IN, EN and Endo cell clusters exhibited the highest average TPS scores, whereas Myeloid cells had the lowest. This pattern suggests that neuronal and vascular components may undergo more pronounced transcriptomic alterations in PD, in contrast to relatively preserved immune-related populations. TPS increased progressively with advancing disease stage (**Supplementary** Fig. 9d); statistical comparison revealed significant elevations in Medium and Late Braak LB groups (t-test, p < 2.3 × 10⁻⁸ and p < 7.6 × 10⁻⁷, respectively) compared to the Control/Early group. In contrast, the difference between Medium and Late stages was modest, indicating that the most substantial transcriptomic alterations occur during the early phases of PD, after which TPS levels plateau, reflecting a stabilization of transcriptional dysregulation in later stages.

To further delineate how different cell types engage disease-associated transcriptional programs across the progression of PD, we ranked cell types within each Braak LB group based on their mean TPS (**Fig. 4c**), enabling a comparative assessment of their relative contribution to transcriptomic pathology at each stage. Mural and endothelial cells ranked highest in later Braak LB groups, supporting their potential role in vascular remodeling and blood-brain barrier dysfunction in PD. In contrast, neurons (EN and IN) exhibited a decline in ranking as disease progressed, consistent with progressive neuronal vulnerability and loss. Interestingly, myeloid cells showed moderate transcriptional activation in early PD stages (Braak LB 0-2) but a diminished response in later stages (Medium, Late). This pattern suggests that early immune activation may transition into a stress-like or senescent state, becoming increasingly disconnected from the dominant transcriptional signatures of advanced PD.

### Dynamic Changes in Cell-Cell Interactions Across PD Progression

Intercellular communication is essential for maintaining brain homeostasis, and its disruption is increasingly recognized as a key feature of neurodegenerative diseases, including PD^52^. However, the molecular mechanisms underlying altered cell-cell signaling in PD remain poorly defined. To characterize the molecular rewiring of intercellular communication in PD, we systematically analyzed ligand-receptor (LR) interaction dynamics across major brain cell classes. Differential interaction analysis comparing PD and controls uncovered a subset of highly dysregulated LR pairs (**Fig. 5a**, **Methods, Supplementary Table 9**), characterized by upregulation of immune- and growth-related interactions, such as FSTL5-CNTN5 and EFNA5-EPHA6, and downregulation of neuronal connectivity-associated pairs, including TNC-CNTN1 and FGF12-FGFR2. Gene ontology (GO) enrichment analysis of the top 5% most dysregulated LR pairs revealed significant overrepresentation of biological processes related to axonogenesis, neuron projection guidance, regulation of nervous system development, ERK1/2 signaling, and epithelial morphogenesis (**Fig. 5b**). These findings suggest a coordinated dysregulation of neurodevelopmental and immune-modulatory signaling pathways in PD.

**Figure 5.**
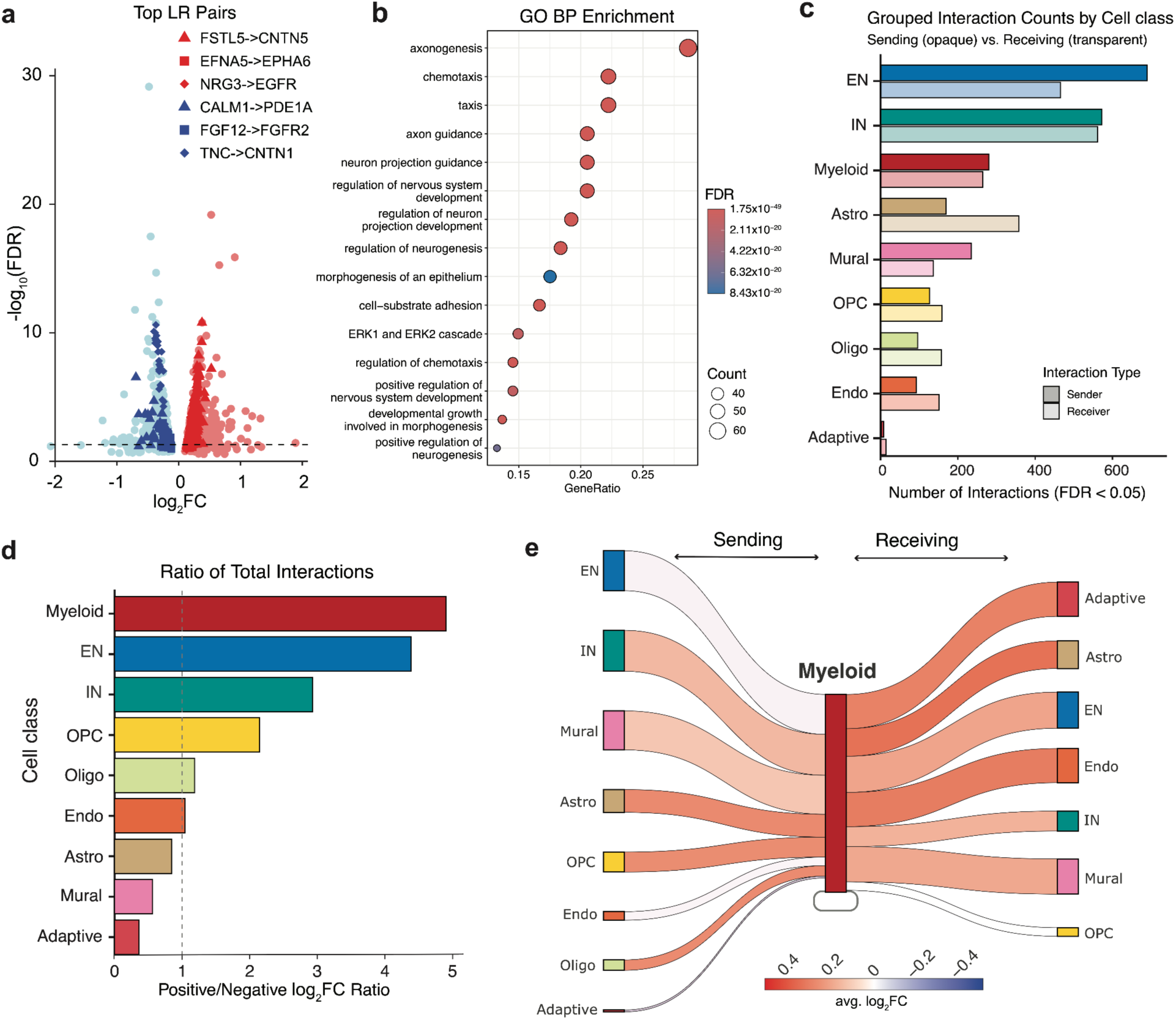
Differential landscape of cell-cell interactions PD. (a) *Differentially regulated LR pairs* ranked by log_2_FC and statistical significance. Top LR interactions are highlighted (red up-regulated, blue down-regulated). (b) *GO Biological Process enrichment analysis* of the top 5% most significantly altered LR pairs reveals overrepresentation of neurodevelopmental and immune-modulatory pathways. (c) *Total number of FDR-significant LR interactions per class*, stratified by sender (opaque) and receiver (transparent) roles. (d) *Log_2_FC ratio of significant up-regulated versus down-regulated CCIs* (FDR < 0.05) across classes. (e) *Sankey diagram depicting the directional flow of LR interactions* centered on Myeloid cells, indicating their dominant role as both signal senders and receivers in PD.

Further examination of the most significantly dysregulated LR pairs (FDR < 0.05, |log_2_FC| > 0.2) revealed distinct biological themes associated with up- and down-regulated interactions in PD (**Supplementary** Fig. 10a-d). Up-regulated LR pairs predominantly converged on immune signaling, lipid metabolism, and developmental pathways (**Supplementary** Fig. 10a-b). Notably, increased APOE-LRP and SPP1-integrin interactions implicate enhanced microglial activation and dysregulated lipid clearance^53–57^. Elevated axon guidance and extracellular matrix remodeling signals, such as SLIT-ROBO, FSTL5-CNTN5, and NTNG1-LRRC4C, further suggest aberrant neurodevelopmental signaling and glial-neuronal crosstalk in the disease state ^58–61^. In contrast, down-regulated LR pairs were enriched for matrix-integrin interactions, guidance molecules (e.g., SLIT3-ROBO1, TNC-CNTN1), and neuroimmune regulators (**Supplementary** Fig. 10c-d), indicating impaired structural maintenance and diminished neuro-glial adhesion^62^. These findings suggest that while immune-related signaling is selectively amplified in PD, pathways supporting tissue integrity, synaptic remodeling, and homeostatic glial support are concurrently suppressed.

Focusing on class level examinations, ENs and INs exhibited the highest number of statistically significant dysregulated interactions (FDR < 0.05), followed by myeloid populations (**Fig. 5c**), implicating neurons and myeloid cells as major hubs of signaling activity. When stratifying interactions by directionality and fold-change polarity, we observed a pronounced shift toward upregulated signaling in myeloid cells, with a positive-to-negative interaction ratio exceeding that of any other class (**Fig. 5d**). Neuronal populations also demonstrated elevated ratios, further supporting a neuro-immune axis of perturbed communication in PD. To further resolve the contribution of immune components, we constructed a directional LR network centered on myeloid cells, revealing extensive bidirectional interactions with multiple neuronal and glial populations (**Fig. 5e**). Notably, the majority of these interactions were upregulated in PD, highlighting a disease-associated amplification of myeloid-mediated signaling. These observations suggest that myeloid cells are prominent mediators of inflammatory and neurodegenerative communication, transducing signals within the broader landscape of cell-cell interactions (CCIs). Their extensive upregulation of ligand-receptor activity in both sending and receiving roles points to a hyperactivated neuroimmune axis as a defining feature of PD pathogenesis.

### Progressive and stage-specific shifts in brain cellular composition in Parkinson’s disease

Understanding how cellular composition shifts across disease stages and clinical variables is critical for disentangling transcriptional changes due to true cellular reprogramming from those driven by shifts in cell-type abundance. To this end, we investigated cell-type compositional changes using crumblr^24^ across multiple conditions; age, Braak LB progression (0-6), diagnosis (PD vs Control) and Braak LB groups early (Braak LB 1-2)/medium (Braak LB 3-4)/late (Braak LB 5-6) (**Fig. 6a**, **Extended Data** Fig. 5, **Methods, Supplementary Table 10**).

**Figure 6.**
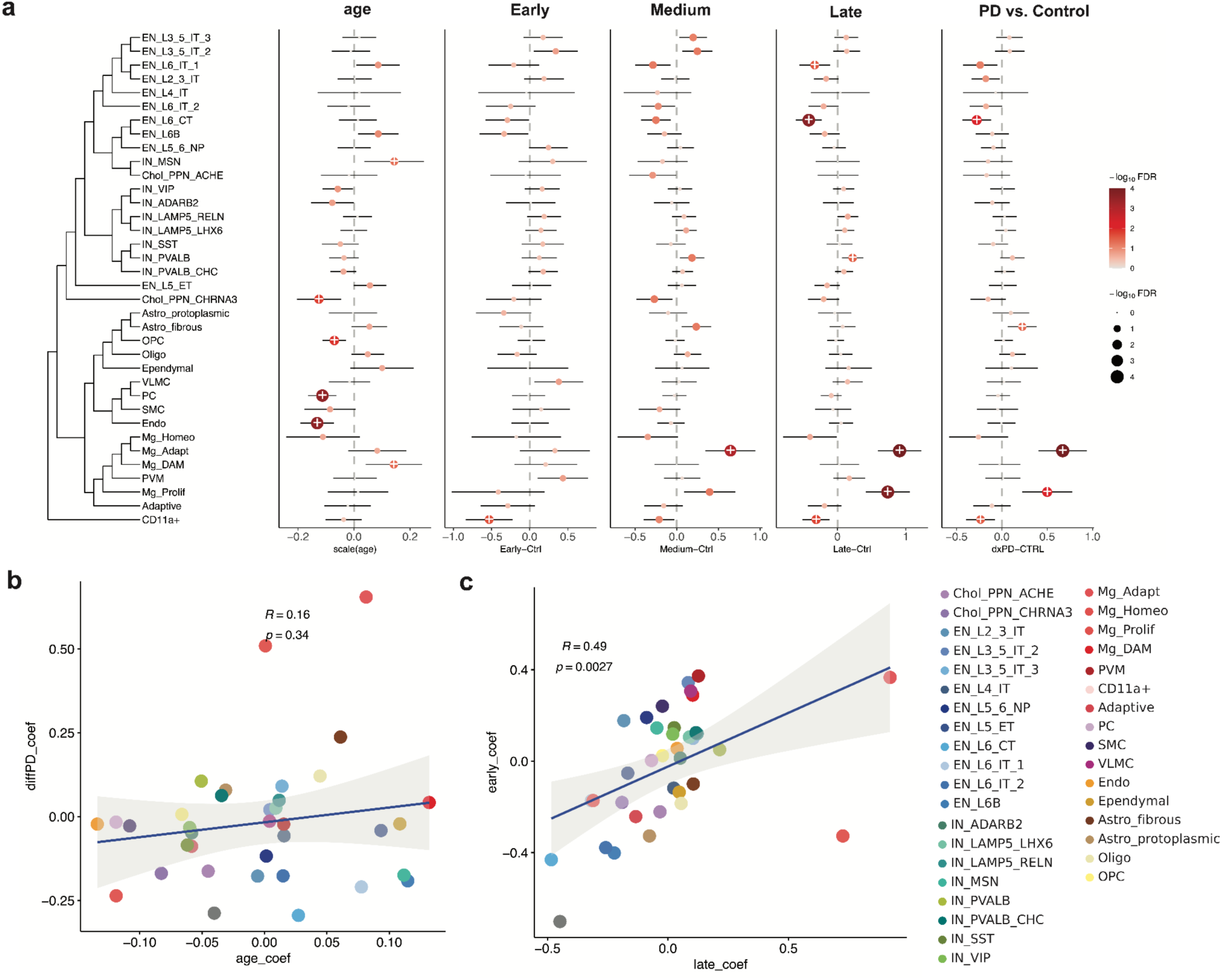
Stage-specific shifts of cellular composition in PD. **(a)** Compositional changes across multiple conditions, comparing early (Braak LB 1-2), medium (Braak LB 3-4), and late (Braak LB 5-6) Braak LB groups against controls, as well as progression along the full spectrum of Braak LB (0-6). Dot size represents effect magnitude, and color indicates significance (-log_10_(FDR)). **(b-c)** Correlation analyses of compositional shifts, showing relationships between cell-type changes across age and disease progression (b), and for early versus late stages of PD (c).

Within neuronal populations, EN_L6_CT and EN_L6B neurons exhibit a consistent decline starting in the early Braak LB group, with progressively greater reductions observed in later stages of PD. In contrast, microglial subclasses displayed divergent trends; homeostatic microglia decreased, whereas adaptive and proliferative microglia expanded, suggesting a shift toward a reactive microglial state as PD progresses.

Interestingly, we observed a subtle increase in the proportion of endothelial and mural cells in the early stages of PD (early group vs. control), potentially reflecting early vascular remodeling or blood-brain barrier (BBB) adaptation. However, unlike the compositional shift, these cell types do not exhibit strong transcriptomic pathology scores (as measured by TPS) until later stages of PD (Braak LB 3-6; **Fig. 4b-c**), suggesting that while their numbers rise early, their disease-associated molecular programs are activated only in more advanced stages of pathology.

Correlation analysis comparing cell-type compositional changes in PD (PD vs. control) with those associated with normal aging revealed a weak correlation (Pearson *r* = 0.16), indicating the distinct cellular remodeling that characterizes PD beyond age-related shifts. Among all cell populations, proliferative and adaptive microglia exhibited the most pronounced divergence between PD and aging, suggesting that PD is associated with fundamentally different microglial activation states and dynamics compared to those observed during typical aging (**Fig. 6b**).

Comparison of cell-type compositional changes between early and late PD groups (**Fig. 6c**), revealed a moderate correlation (Pearson *r* = 0.49), suggesting partial degree of continuity in cellar remodeling with PD progression. However, specific myeloid subclasses, including proliferative microglia and PVMs, displayed stage-specific patterns, indicating that distinct immune programs may be differentially engaged across different phases of PD pathology.

Finally, we validated our cell-type compositional findings by repeating the analysis separately in each brain region (**Extended Data** Fig. 5a-c). We observed strong correlations in cell-type compositional shifts across cortical regions, whereas subcortical regions showed lower concordance. In particular, the DMNX region exhibited the highest variability in compositional changes (**Extended Data** Fig. 5a-c), which may reflect increased biological heterogeneity and different cellular representations (such as the absence of EN, IN neurons).

To further validate the robustness of our findings, we assessed cell-type compositional changes in an independent cohort by analyzing PD cases and matched controls from the PsychAD dataset^20,63^ (**Extended Data** Fig. 5g-h). At the cell class level, compositional shifts associated with PD diagnosis (PD vs. control) showed a moderate correlation with the discovery cohort (Pearson *r* = 0.51, *p* = 0.16), while changes associated with age progression demonstrated a strong correlation (Pearson *r* = 0.79, *p* = 0.011). These results support the reproducibility of PD-related cellular alterations across cohorts and reinforce the biological relevance of the observed compositional dynamics.

### Myeloid subtypes exhibit distinct responses across different stages of PD

Microglia are among the earliest responders to neurodegenerative pathology and are increasingly recognized as key modulators of inflammation and neurodegeneration in PD^64^. However, the diversity and stage-specific dynamics of myeloid cell subtypes in the human PD brain remain incompletely characterized. Given this, we performed an in-depth analysis of the myeloid compartment across the PD trajectory. We extracted myeloid nuclei from the full AMP-PD single-nucleus dataset, isolating 118,515 nuclei annotated as microglia, PVMs, and CD11a+ (monocyte-derived vascular-associated cells)^65,66^ populations. Using a clustering-based approach combined with reference-guided annotation^67^ (**Methods**), we identified 6 subclasses and 13 distinct transcriptional subtypes (**Fig. 7a**). Each subtype was associated with robust marker gene signatures (**Fig. 7b**), consistent with previous studies^67,68^ (**Extended Data** Fig. 6a-d), and exhibited distinct transcriptomic phenotypes (**Extended Data** Fig. 6e).

**Figure 7.**
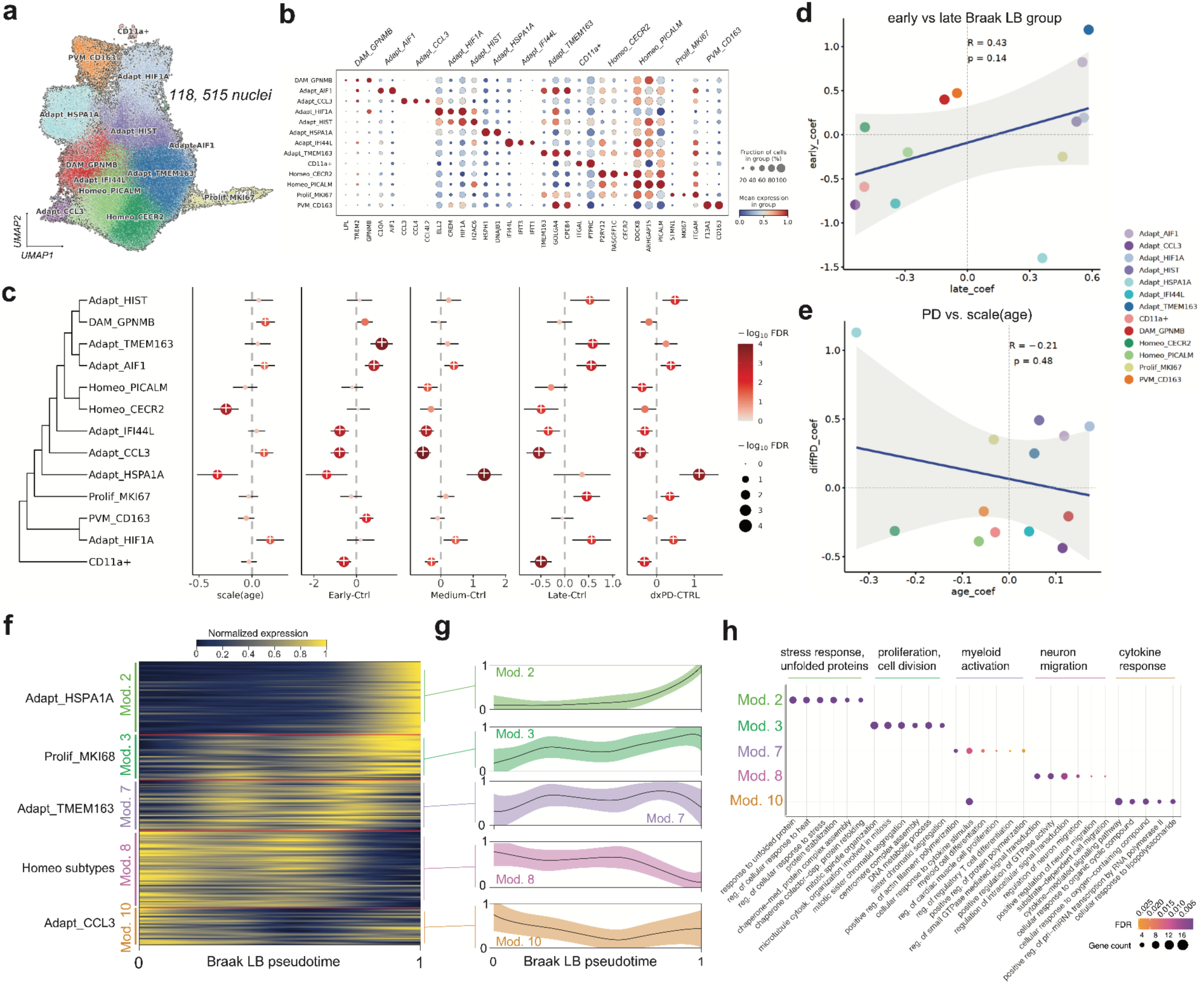
Functional Remodeling and Subtype-Specific Responses of Myeloid Cells Across PD Progression. **(a)** *UMAP embedding of myeloid nuclei* (n = 118,515) revealing distinct transcriptional subtypes including homeostatic, adaptive, proliferative microglia, PVMs, and CD11a⁺ populations. **(b)** *Expression of key marker genes for each myeloid subtype*, based on reference markers curated from Lee et al. (2023)^67^. **(c)** *Cell-type compositional changes across disease progression using crumblr analysis*^24^. Subtype-level comparisons shown for age progression, early (Braak 1–2 vs. 0), medium (Braak 3–4 vs. 0), late (Braak 5– 6 vs. 0), and PD vs. control contrasts. Dot size and color reflects the -log_10_(FDR), and the x-axis position represents the effect size (log_2_FC) of compositional changes. **(d)** *Comparison of subtype-level compositional shifts* between early and late Braak LB stages (Pearson *r* = 0.43, *p* = 0.14). **(e)** *Scatter plot comparing PD-related compositional shifts* (PD vs. control) with natural aging trends, highlighting subtype-specific divergence (Pearson *r* = -0.21, *p* = 0.48). **(f)** *Expression heatmap of genes from selected hotspot modules* (2, 3, 7, 8, 10), capturing subtype-specific responses along inferred Braak LB pseudotime. **(g)** *Smoothed trend lines of average expression for modules* shown in panel (f) across pseudotime. **(h)** *GO enrichment of biological processes* for hotspot modules. Dot color indicates FDR significance, and size reflects the number of genes contributing to each term.

To assess the functional diversity within the myeloid population, we applied a module-based transcriptomic analysis using Hotspot^69^ (**Methods**), providing an orthogonal perspective to cluster-based subtype assignments. The top 1,000 highly variable genes across all myeloid cells were grouped into 11 co-expression modules (**Extended Data** Fig. 6f-g**, Supplementary Table 11**), each associated with distinct biological functions such as inflammatory activation, stress response, phagocytosis, and proliferation (**Extended Data** Fig. 6h). Derived Hotspot modules showed substantial agreement with previous observations of myeloid function in neurodegeneration^67^.

To link functional profiles with cell-type compositional changes during PD progression, we repeated the crumblr^24^ compositional analysis at the level of individual myeloid subtypes (**Fig. 7c-e, Supplementary Table 12**). This high-resolution analysis broadly recapitulated trends observed in broader cell-class analyses (**Fig. 6a**), while revealing divergent subtype-specific behaviors across Braak LB stages (**Supplementary** Fig. 11). Homeostatic microglia remained relatively stable in early PD, but their decline became fully apparent in mid- to late-stage disease, suggesting a breakdown of their homeostatic surveillance role. In contrast, adaptive subtypes such as Adapt_TMEM163 and Adapt_AIF1 were sharply upregulated in early PD, and this elevated state was maintained through later stages. Conversely, other adaptive populations, including Adapt_IFI44L and Adapt_CCL3, displayed early downregulation and remained suppressed throughout the disease course.

Notably, the Adapt_HSPA1A subtype, associated with stress responses and unfolded protein detection, showed a biphasic pattern -downregulated in early PD but upregulated in mid- to late stages. This pattern reflects a delayed engagement of proteostasis mechanisms and a potential shift toward a reactive or senescence-like microglial state in response to chronic stress. A similar pattern was observed for the Prolif_MKI67 subtype, which may reflect a late-stage proliferative or reactive transition in the microglial population. Interestingly, both PVM_CD163 and disease associated microglia (DAM) exhibited a biphasic transition, however with the opposite trend, showing a modest increase in early PD followed by a subtle decline in later stages. Interestingly, the behaviour of DAM in PD is distinct from that observed in AD associated microglia (ADAM)^67^.

Comparison with aging-related compositional trends (**Fig. 7c first panel, Fig. 7e**) revealed that several subtypes exhibited distinct or inverted dynamics. Specifically, the Prolif_MKI67, DAM, and Adapt_HSPA1A subtypes followed similar trajectories in early PD and aging, but reversed their patterns in mid-to-late PD, suggesting a shift toward disease-specific regulatory states. In contrast, Adapt_TMEM163, Adapt_AIF1, Adapt_HIF1A, and homeostatic microglia maintained compositional trends similar to those seen in aging, indicating possible shared programs. Meanwhile, Adapt_IFI44L and Adapt_CCL3 displayed opposing trends in PD vs. age progression, suggesting a disease-specific suppression of certain inflammatory states.

To further resolve the transcriptional programs underlying myeloid compositional shifts in PD, we applied a neural network-based predictive modeling framework to infer a continuous Braak LB progression score, referred to as Braak LB pseudotime, for individual nuclei, based on their gene expression profiles (**Extended Data** Fig. 7a-c**, Supplementary** Fig. 12a**; Supplementary Table 13; see Methods**). We then leveraged this model to trace expression dynamics of selected Hotspot modules across representative myeloid subtypes (**Fig. 7f-g**). This enabled us to classify myeloid subtypes based on their engagement with distinct functional programs across inferred Braak LB progression (**Fig. 7h**). Specifically, stress response and proteostasis-related programs, including response to unfolded proteins (Mod. 2), show a marked increase in late stages of predicted Braak LB. Proliferation and cell division-related genes (Mod. 3) begin to rise in early stages and continue to increase progressively along the predicted Braak LB trajectory. In contrast, myeloid activation (Mod. 7) is elevated early in the disease and remains relatively stable with minor fluctuations. Genes associated with neuronal migration and signal transduction progressively decline (Mod. 8), while cytokine signaling is activated early, decreases in mid-stage PD, and shows a mild resurgence in later stages (Mod. 10).

Furthermore, we compared disease progression trajectories of selected hotspot modules between PD and AD (from the PsychAD dataset^20^) (**Supplementary** Fig. 12b**, Supplementary Table 14; see Methods**). Overall, we observed a similar trend in the expression of proliferation (Mod. 3), myeloid activation (Mod. 7), and cytokine production (Mod. 10) modules between PD and AD microglia, indicating shared aspects of immune activation and cell cycle regulation across diseases. In contrast, the stress response (Mod. 2) and neuronal migration (Mod. 8) modules displayed diverging dynamics, with the stress response module in AD exhibiting a markedly lower and flat expression profile along the Braak pseudotime. This suggests distinct, disease-specific regulation of cellular stress mechanisms in PD compared to AD.

## Discussion

PD has long been characterized by the degeneration of dopaminergic neurons in the substantia nigra, with α-synuclein aggregation and Lewy body pathology serving as its histopathological hallmark^1^. However, growing evidence from neuropathological, transcriptomic, and imaging studies suggests PD is a multisystem disorder, involving diverse brain regions and multiple non-neuronal cell types, including microglia, astrocytes, oligodendrocytes, and vascular cells^4–6,8,9^. Previous single-cell studies have provided essential insights into selective neuronal vulnerability in the substantia nigra^10,11^ and inflammatory activation in glial cells^5,64^, but most have been limited to a single brain region, late-stage disease, or restricted sample sizes.

Here, we provide comprehensive analyses of the first large-scale, multi-regional single-nucleus transcriptomic atlas of PD, integrating molecular, genetic, and cellular features across nearly 100 donors and five anatomically distinct brain regions^16^. While regional differences were modest at the transcriptomic level, our findings converge on a shared architecture of transcriptional remodeling that unfolds in a temporally structured, cell-type-specific manner, forming the basis for a refined conceptual model of PD progression.

A central feature of this model is a temporal cascade of transcriptional changes that begins in microglia and neurons and culminates in vascular cell engagement. In early disease stages, we observe activation of adaptive microglial programs, including lipid clearance and proteostasis responses, extending prior observations of early microglial reactivity and α-synuclein phagocytosis^5,64,72^. As disease advances, these programs shift toward proliferative and stress-associated states, including signatures of oxidative stress and cell cycle re-entry. These transitions are largely absent in normal aging, emphasizing their disease specificity. Similarly, excitatory and inhibitory neurons display progressive upregulation of unfolded protein response (UPR) and DNA damage pathways, consistent with prior reports linking proteostasis failure to α-synuclein toxicity^73,74^ ^75–77^, but here contextualized with high resolution across cell types and disease stages.

Vascular populations, including endothelial and mural cells, do not show major transcriptional changes early on, but become increasingly engaged in morphogenesis, barrier function, and stress-related pathways at later stages. These findings align with prior reports of blood-brain barrier dysfunction in PD^9,78^ but now clarify the temporal delay between compositional shifts and transcriptional activation, suggesting that vascular dysregulation may be a consequence, rather than a driver, of early pathology.

Importantly, we show that these transcriptional dynamics are shaped by genetic risk. Integration with PD GWAS data and TF-target gene networks reveals convergence on key transcriptional regulators (HSF1, FOXO3, and MYC) that are upregulated across multiple lineages and implicated in proteostasis, oxidative stress resilience, and immune modulation^38,40,79–83^. These regulators are potential mechanistic links between inherited susceptibility and downstream transcriptomic pathology. The TPS quantifies molecular burden per cell type and individual, revealing early burden in neurons and microglia, and later engagement of vascular cells, providing a scalable framework for staging molecular pathology.

In summary, our findings support a modular model of PD pathogenesis, where genetically encoded risk factors influence the magnitude and timing of transcriptional stress responses across a multicellular brain ecosystem. This model builds on prior literature suggesting non-neuronal contributions to PD, but provides new granularity, defining cell-type–specific trajectories and regulatory circuits in human brain tissue across disease stages. Rather than viewing PD as primarily a neuron-intrinsic disorder, our data suggest it is a progressive failure of intercellular stress adaptation, marked by early immune compensation, neuronal vulnerability, and delayed vascular disruption.

Future efforts should combine spatial transcriptomics, *in situ* hybridization, and functional perturbation to localize and validate the molecular events identified here. This atlas lays a foundation for uncovering stage-specific therapeutic targets, guiding biomarker development and the rational design of interventions aimed at halting or reversing the cellular cascade of PD.

## Methods

### Cohort data collection

The AMP-PD cohort consists of genetic (WGS) and transcriptomic (snRNA-seq) data from 100 donors sourced from the following brain banks: NIH NeuroBioBank at the Mount Sinai School of Medicine, NIH NeuroBioBank at the Harvard Brain Tissue Resource Center, NIH NeuroBioBank at the University of Miami, the University of Miami Brain Endowment Bank and the University of Miami Udall Center of Excellence for Parkinson’s Disease Research (**Fig. 1**, **Extended Data** Fig. 1, **Supplementary Table 1**). Raw data described in this study are available for use by the research community and have been deposited in the AMP PD Knowledge Platform^84^; select “AMP PD Release 4”).

All data were obtained from biobanks with appropriate informed consent from all participants. Detailed cognitive, neuropathological, and demographic information was gathered for all donors, who were mainly of European descent with a male-to-female ratio of 3:2 (**Extended Data** Fig. 1, **Supplementary Table 1**). The study included 75 clinically diagnosed PD cases and 25 neurotypical controls (**Supplementary Table 1**). Additionally, data were collected on Braak NFT staging as a measure of co-occurring Alzheimer’s pathology^19^, which evaluates tau neurofibrillary tangle accumulation, and the Hoehn and Yahr scale^85^, assessing functional disability in PD. The importance of examining both neuropathological (Braak LB staging) and detailed clinical characteristics of PD is evident from only a limited correlation observed between those phenotypes (**Fig. 4a-b, Supplementary** Fig. 9a). Furthermore, the availability of both Braak AD and Braak PD stages provides an opportunity to explore the transcriptomics basis of symptomatic, clinical^86^, and, to a lesser extent, genetic overlaps^87,88^ between Tau and Lewy body accumulation. However, it is important to note that all donors were either unaffected controls or clinically diagnosed exclusively with PD but no other neurological or major neuropsychiatric diseases, including AD.

### snRNA-seq data generation and preprocessing

#### Nuclei isolation and snRNA-seq library preparation

Sample processing, including nuclei isolation and snRNA-seq library preparation, was described previously^16^.

#### Computational preprocessing

Sequencing alignment, genotype-based demultiplexing, and donor assignment were performed as previously described^16^. Briefly, reads were aligned to the hg38 genome using STARsolo (v2.7.9a)^89^, and cell assignments were made using cellSNP-lite (v1.2.0)^90^ and vireo (v0.5.8)^91^, with genotype concordance verified via QTLtools-mbv (v1.3)^92^. Cells failing basic QC criteria, including low gene/UMI counts, high mitochondrial content, or genotype discordance, were excluded. A three-tiered quality control pipeline was implemented to remove ambient RNA contamination, low-quality cells, and doublets (identified with Scrublet^93^ v0.2.3). Genes expressed in fewer than 0.05% of cells and samples with fewer than 50 nuclei were also filtered. These steps ensured a high-confidence dataset suitable for downstream analyses.

#### Cell clustering

A total of 2,092,520 nuclei passing the initial QC steps were consolidated into a single dataset for downstream analysis. Normalization and clustering were performed using SCANPY (v.1.9.3)^94^ and the Pegasus package (v1.10.1)^95^. Briefly, nuclear counts were scaled by total library size and log-transformed. To identify highly variable genes, 6,000 genes with the greatest dispersion and mean were selected, excluding genes on sex chromosomes and mitochondria-related genes. Technical effects from library size, percentage of mitotic reads, and cell cycle variations were regressed out using the *pg.regress_out()* function in Pegasus. Batch effects arising from the use of different brain banks were addressed with the Harmony algorithm via the *pg.run_harmony()* function.

Dimensionality reduction was performed by first applying Principal Component Analysis (PCA) on the variable genes, followed by Uniform Manifold Approximation and Projection (UMAP) using the top 30 principal components (PCs). It was confirmed that more than 30 PCs accounted for 100% of the data variance. The top 50 PCs were subsequently used to construct a k-nearest neighbors (k-NN) graph with k=100, and the Leiden algorithm was applied to identify clusters. These steps were executed using Pegasus functions *pg.pca()*, *pg.elbowplot(), pg.neighbours()*, and the Leiden clustering tool *sc.tl.leiden()* from SCANPY.

Differential gene expression for each cluster was analyzed using the variance-adjusted t-test implemented in SCANPY’s *sc.tl.rank_genes_groups()* function. The top 300 genes per cluster were identified and tested for overlap with previously established markers. During iterative sub-clustering, potentially spurious clusters were flagged and excluded. These included clusters displaying extreme separation within sub-cluster populations, unusually high total counts, or mixed expression of markers from multiple cell types, indicative of doublets or low-quality nuclei. This stringent process ultimately yielded 2,092,520 nuclei for downstream analyses.

Class-level cellular identities were validated by calculating cosine similarity correlations between the pseudo-bulk transcriptomes of Leiden clusters and reference datasets (Siletti et al. (2023)^26^, Mathys et al. (2023)^21^, Lee et al. (2024)^20^), as well as in depth marker gene examination. This ensured robust alignment with established cell type classifications. Furthermore, for myeloid subtype classification, we utilized microglial subtypes defined in Lee et al. (2023)^67^.

### Variance Partition Analysis of Gene Expression

To analyze variance in gene expression, pseudobulk data were aggregated by library and stacked across cell types using the *StackedAssay* function in dreamlet^23^. This approach enabled a comprehensive analysis spanning multiple cell types. Variance partitioning^28^ was conducted on the resulting stacked pseudobulk dataset using the following regression model:

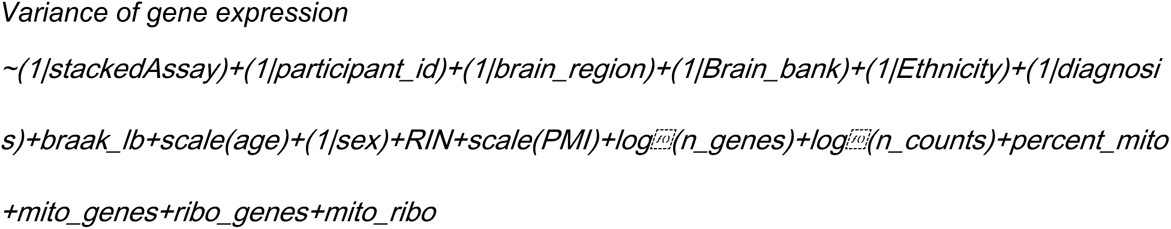

Here, *diagnosis* represents the binary disease status for PD, while *braak_lb* accounts for Braak LB staging, and additional terms include both biological (e.g., age, sex, ethnicity) and technical covariates (e.g., RIN, PMI, mitochondrial content).

### Compositional variation analysis using Crumblr

To assess variation in cell type composition, we utilized the crumblr method^24^. Briefly, crumblr applies a centered log-ratio (CLR) transformation to scale cell count ratios (i.e., fractions) and incorporates these into linear models. However, CLR-transformed data remains highly heteroskedastic, resulting in widely varying measurement precision. To address this, crumblr employs a fast asymptotic normal approximation of CLR-transformed counts derived from a Dirichlet-multinomial distribution. This approach models the sampling variance of the transformed counts and enables the incorporation of this variance as precision weights into linear (or mixed) models. These precision weights enhance statistical power while maintaining control over the false positive rate.

Additionally, crumblr applies a variance-stabilizing transformation based on precision weights, improving the performance of downstream analyses such as PCA and clustering. Hypothesis testing was performed using the following formulae (based on prioritization of covariates using variance partitioning^28^ approach on cell type composition object):

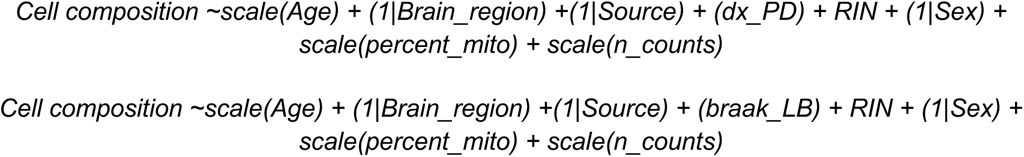

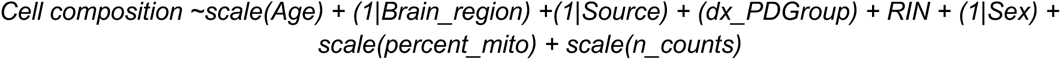

Where *dx_PD* represents PD case-control status as a binary variable; *braak_lb* presents Braak LB values; *dx_PDGroup* represents Braak LB group (Early, Medium, Late)*; Source* represents brain bank and *RIN* is RNA integrity number. In these models, categorical covariates were specified as random effects, while continuous variables were included as fixed effects. For categorical variables of interest, the model was parameterized with a zero intercept, and predefined contrasts were applied to directly compare the relevant factor levels.

### Differential gene expression analysis using Dreamlet

Analyzing millions of single cells across diverse phenotypes in a large-scale disease atlas poses significant computational challenges^96^ and can be suboptimal using standard single-cell approaches^97–100^. To manage the complexity of such data, including repeated measures and low read counts per cell, we utilized *dreamlet*^23^ for differential expression analysis.

Dreamlet builds on the statistical framework of *dream*^101^, leveraging a pseudobulk approach^97,98102^ to overcome these limitations. By aggregating cells at the donor level, it reduces noise and allows the application of linear mixed models for single-cell omics data. For each feature and cell cluster, the following mixed models were applied:

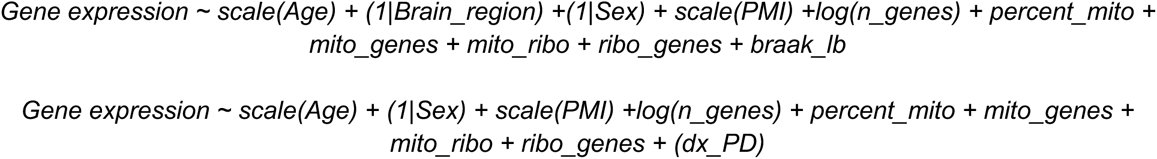

Where *dx_PD* is represented as a contrast for *PD vs. control*. In this model, categorical variables were treated as random effects, while numerical variables were modeled as fixed effects. For categorical phenotypes of interest, the intercept was set to zero, and pre-defined contrasts were applied to compare the two factors.

### Gene Set Enrichment Analysis (GSEA)

Gene set enrichment analysis was run using package enrichR (v3.4)^30^ and clusterProfiler (v4.16.0)^103^ with default parameters. Gene set enrichment was calculated with the following libraries: GO_Biological_Process_2023, WikiPathway_2023_Human, Reactome_2022.

To enhance the interpretability of differential expression (DE) results, we categorized 180 unique GO biological processes (FDR <0.05) into a coherent and non-redundant set of meta-functional terms based on semantic similarity. This grouping was performed using the rrvgo (v1.20.0)^31^ package, which clusters functionally related GO terms into broader biological themes. This approach resulted in the identification of 19 distinct and interpretable functional themes, providing a streamlined representation of enriched biological processes.

### Hotspot module analysis (HSA)

#### Identify microglial gene expression modules

The normalized AnnData object containing only myeloid populations (microglia, PMV and CD11a+) was used as the input for Hotspot (v0.9.0)^69^, with latent_obsm_key = ‘X_pca’ and model = ‘normal’. Gene modules were called on the Hotspot object after creating an KNN graph (n_neighbors = 30) and using top 1,000 hvg, computing autocorrelations and local correlations, using the create_module function with min_gene_threshold = 50 and fdr_threshold = 0.05. A total of 12 modules were returned as output.

### Transcription Factor (TF) Inference

To identify key transcriptional regulators underlying differential gene expression, we inferred TF activity using the univariate linear model (*ulm*) method implemented in the decoupleR package (v1.9.2)^48^. This method fits a linear model for each TF, predicting observed gene expression based on its TF-Gene interaction weights. The resulting t-value of the slope serves as the TF enrichment score, where positive scores indicate activation and negative scores indicate inhibition.

We leveraged CollecTRI^49^, a curated regulatory network integrating TF-target interactions from 12 different resources, providing increased TF coverage and enhanced sensitivity compared to DoRothEA^104^ and other literature-based gene regulatory networks. Interactions in CollecTRI are weighted by their mode of regulation (activation/inhibition) to improve biological relevance. TF inference was conducted on our DEG results from dreamlet for contrast *PD vs. control*, allowing us to identify dysregulated TFs associated with PD pathology. TF-TF connectivity was assessed using STRING database^50^ (https://string-db.org/) for protein-protein interaction (PPI) predictions.

From the top upregulated, cell type-specific TFs (**Fig. 3b**), we derived TF-target gene networks and systematically evaluated their overlap with GWAS-prioritized genes by varying the number of included targets (**Supplementary** Fig. 7, **Supplementary** Fig. 8b-c).

### Transcription Pathology Score (TPS)

To quantify PD-associated transcriptional changes at the individual subject level, beyond traditional binary case-control DE analysis, we adopted a more continuous measure of transcriptional pathology based on cell-type-specific pseudobulk profiles.

For each cell type, we first calculated the mean gene expression pattern across all subjects. Next, we computed the partial Pearson’s correlation between each subject’s expression profile and the *PD vs. control* differential expression profile obtained from our joint-brain region DEGs using dreamlet, while adjusting for the baseline (mean) expression of cell-type specific genes within the given cell type. This approach generated an individual-by-cell type Transcriptomic Pathology Score (TPS) matrix, capturing the extent of transcriptional dysregulation per subject. Finally, to obtain a donor-level TPS, we averaged TPS values across all cell types for each individual, providing a comprehensive measure of transcriptional pathology in PD. Cell-type specific TPS was calculated only for cell types which were sufficiently enriched in all donors.

### Trajectory analysis using neural network models

#### Rationale

Traditional linear models have been instrumental in identifying gene expression changes associated with disease. However, gene regulation is often nonlinear, shaped by complex interactions and feedback loops that linear models may not fully capture. Pseudotime methods (such as Monocle3) aim to model such dynamics by identifying continuous transitions in gene expression. However, disease-related variance is often overshadowed by confounding factors like donor identity, sex, and brain region. When disease signals are not the dominant source of variation, trajectory methods may instead reflect these confounders. To address this, we previously trained neural network models that directly infer disease progression from gene expression^20^. This supervised approach enables us to assign each cell a continuous disease score while explicitly modeling its relationship to gene expression, allowing us to isolate subtle disease-specific signals despite substantial background variability. Here, we follow a similar approach to model how gene expression varies with the Braak stage on a fine resolution timescale.

#### Model architecture

The neural network model was a relatively simple feedforward network with three hidden layers. The input to the neural network were the log1p transformed gene counts from individual cells. Each hidden layer consisted of 1,024 units using the ReLU activation function. Layer and batch normalization were not used. The output of the model was the Braak LB stage prediction.

#### Model training

Single-cell gene counts from the top 8,000 protein coding genes for each cell class, based on the percentage of cells the gene was expressed in, were used for training. We found that overfitting became more problematic when using a greater number of genes (data not shown). Genes found on sex chromosomes were excluded to discourage the model from learning to associate the Braak LB stage status with sex.

When training the model to predict the Braak LB stage (as a proxy for PD pathology), target values were first normalized to zero mean and unit standard deviation, and the mean-squared error loss was used. Both loss terms were trained simultaneously. To prevent the model from overfitting the data, we applied dropout with probability of 0.5 to all hidden layers (after the ReLU activation). For each cell class, we divided the cells into 20 splits. In each split, ∼95% of the cells were used for training, and the remaining ∼5% were used for inference. Within each split, cells from a single donor exclusively belonged to either the training set or the inference set, but never both. Thus, model predictions were always based on cross-validated data from different donors. We trained one model for each of the 20 splits to generate predictions for all donors for that cell class.

For each split, we trained for 30 epochs, and then averaged the predicted Braak scores of each cell across the last five epochs. The models were trained using stochastic gradient descent (SGD) without momentum and with a learning rate of 0.02. We found that training with SGD led to greater accuracy compared to adaptive optimizers such as ADAM (data not shown). We used a batch size of 256, and linearly increased the learning rate across 2,000 training steps until it reached 0.02. Finally, we clipped the gradient norm to 1.0 to stabilize training.

#### Calculating gene trajectories

We wished to measure how gene expression varied as a function of the predicted Braak stage. First, gene counts were normalized so that each cell’s total count was 10,000, followed by the log1p transformation. Second, for each donor *j*, we calculated the mean predicted Braak stage *b_j_*, and the mean normalized expression for each gene *y_j,i_*, where the index *i* is over genes. Braak scores and gene expression were pooled across all five brain averages when calculating the means. Averaging within each donor reduced variability and ensured that donors with greater cell counts did not contribute disproportionately in downstream analysis.

Finally, we computed smoothed gene trajectories, *f_i_*(*x*) across predicted Braak by smoothing the expressions with a Gaussian kernel

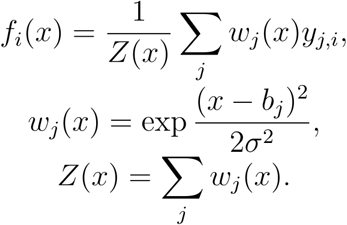

We found σ^2^ = 0.1 provided a good balance between sufficient smoothing without losing temporal specificity. *x* ranged between the minimum and maximum donor-averaged predicted Braak scores, equally divided into 1000 steps.

#### Calculating hotspot trajectories

Given the smoothed gene trajectories defined above, we calculated normalized gene trajectories for genes in hotspot modules. Specifically, each gene trajectory *f_i_*(*x*), where *i* runs across genes and *x* across predicted Braak, was normalized between 0 and 1. We found that different normalization strategies did not qualitatively affect the results (data not shown).

#### Identifying trajectory transition points

To determine whether specific stages of disease progression are associated with shifts in gene expression dynamics, we first projected the normalized gene expression matrix *y_j,i_* onto a lower-dimensional latent space using Partial Least Squares (PLS) regression. The model was trained to best capture variation in donor-averaged predicted Braak LB scores, using 10 components and no feature scaling. Visualization of the resulting projection (**Extended Data** Fig. 7c) revealed at least one apparent nonlinear transition as PD progressed.

To localize these transitions, we smoothed gene expression trajectories and projected them into the PLS space. We then computed the directional derivatives of the trajectories across the range of predicted Braak LB scores, capturing how the overall direction of gene expression changes with disease progression. Using KMeans clustering (with k=3) on these directional vectors across Braak scores revealed two transitions, indicated by the cyan and magenta circles in **Extended Data** Fig. 7c.

While the choice of three clusters was heuristic, the resulting transition points aligned well with independent signals: (1) changes in the relative abundance of microglial subtypes across Braak scores (**Extended Data** Fig. 7d, dashed vertical lines), and (2) shifts in pathway enrichment dynamics (**Extended Data** Fig. 7e and **Supplementary** Fig. 12a, as described below.

#### Pathway enrichment dynamics

To further characterize how the immune response evolves during disease progression, we performed pathway enrichment using a sliding window analysis.

We calculated the change in pathway expression using the smoothed gene trajectories, *f_i_*(*x*), where each window covered 5% of the trajectory, and incrementing by 2% at each interval. The Zenith^105^ z-scores were calculated for each window. We only included pathways for which the Zenith FDRs were below 0.001 for at least 4 windows, and included all pathways with between 10 and 250 genes. Since we were only interested in pathways that could be informative of the mechanisms underlying PD progression, we excluded pathways containing words referring to overly broad behaviors or cognitive functions (“learning”, “memory”, “vocalization”, “social”, “auditory”, “startle response”, “behavior”, “locomotor”, “startle”, “prepulse inhibition”), terms referring to anatomical structures other than the cortex (“substantia nigra development”, “cardiac”, “coronary”, “aortic”, “ventricular”, “kidney”, “metanephric”, “retina”, “optic”, “bone”, “respiratory”, “pulmonary”, “olfactory”, “sperm”, “placenta”, “egg”, “embryonic”, “ovulation”, “estrous”, “placenta”, “sperm”, “mamary”, “germ layer”, “outflow tract septum”, “adrenal”, “epithelial”, “skeletal”, “otic”, “head”) or overly broad neural terms (“nervous system process”, “cerebral cortex”, “recognition”, “host”, “organ”, “developmental growth”).

The full list of significant pathways is shown in **Extended Data** Fig. 7a. We performed hierarchical clustering, based on the Zenith z-score dynamics, to divide the pathways into 12 broad clusters, indicated by the pathway font color. We then selected several representative pathways from each cluster for **Extended Data** Fig. 7e.

#### Comparing hotspot trajectories between Parkinson’s and Alzheimer’s Diseases

To compare gene expression dynamics between AD and PD, we trained neural networks as described above using the PsychAD dataset^20^, which includes both AD and control cases. This allowed us to generate AD gene expression trajectories as a function of the normalized predicted Braak stage. We then computed hotspot scores for AD using the same approach as previously applied to PD.^20^ (**Supplementary** Fig. 12b).

### Inference of Cell-Cell Interactions

Cell-cell interaction (CCI) analysis infers ligand-receptor (LR) interactions based on a gene expression matrix with annotated cell types. By referencing a curated LR resource, CCIs can be inferred and scored using various computational methods. LR pairs between cell types that meet an expression threshold and exhibit differential co-expression are identified as putative interactions.

For this analysis, we used the LIANA framework (v1.4.0)^25^, which integrates multiple established CCI inference methods (CellPhoneDB, Connectome, log2FC, NATMI, SingleCellSignalR, cell2cell package and CellChat)^106–112,113,114^. LIANA aggregates results from these methods using the RobustRankAggregate algorithm^25,113^, generating a consensus magnitude score for each inferred interaction. We applied LIANA’s standard rank_aggregate pipeline (resource_name = ‘consensus’, expr_prop = 0.1, min_cells = 5, n_perms = 1000) separately for each donor in the AMP-PD cohort. To qualify for CCI scoring, each LR gene pair had to be present in the reference resource, expressed in at least 10% of each interacting cell type, and detected in a minimum of 5 cells.

To assess differentially expressed CCIs in PD, we applied the dream linear mixed model^101^, incorporating relevant biological and technical covariates, consistent with our prior analyses of compositional variation and differential gene expression. The model was fitted to interaction-level estimates across two measures of disease progression: binary diagnosis (PD vs. control) and continuous Braak LB staging.

To interpret the biological significance of PD-associated CCIs, we conducted GSEA using the enrichR, clusterProfiler and the Human Gene Ontology Biological Processes 2023 reference^115^ as described previously. CCIs were aggregated by sender and receiver subtypes to improve statistical power, and normalized enrichment scores were computed for each biological process based on the PD-relevant LR gene set. We prioritized pathways that passed the FDR < 0.05 threshold (**Fig. 5b**).

### Genetic correlation and integrative analysis

We examined the convergence between genetic risk and transcriptional dysregulation in PD by integrating GWAS findings with differential expression profiles obtained from dreamlet analysis (see **Methods**). Protein-coding genes nominated through QTL mapping in the original PD GWAS^34^ formed the primary set for analysis. For loci lacking QTL-based nominations, gene assignments were supplemented using annotations from the NHGRI-EBI GWAS Catalog, which prioritizes candidate genes based on their proximity to lead SNPs and additional functional evidence.

### Single-cell polygenic disease risk score

We applied the scDRS package (v1.0.1)^22^ to evaluate the combined expression of disease-associated genes identified through GWAS summary statistics using MAGMA^116^. Each gene’s contribution was weighted by its corresponding MAGMA z-score and inversely weighted by its gene-specific technical noise level in the single-cell dataset. The analysis was performed across all nuclei from the AMP-PD snRNA-seq dataset to compute raw disease scores for each nuclei. Additionally, 200 sets of matched control scores were generated, with control gene sets matched to the disease gene sets based on size, mean expression, and expression variance. Both raw disease scores and control scores were subsequently normalized within each cell to produce normalized disease scores.

These calculations were executed using the *scdrs.score_cell* function, with the following parameters:

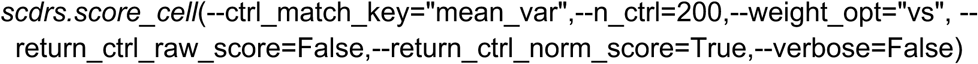

For downstream analysis, we aggregated cell-level scores at the subclass level to assess disease associations across predefined taxonomic subclasses, enabling the evaluation of heterogeneity in disease relevance across distinct cellular populations. This step was performed using the *scdrs.method.downstream_group_analysis* function with default settings. The full results of the subclass-level analysis, including trait information are provided in **Supplementary Table 3**.

### Rare variants

We assessed the burden of rare PD-associated variants by intersecting our WGS dataset with a list of 679 rare SNPs reported in the largest rare variant meta-analysis to date^117^. Of these, 72 SNPs were present in our cohort. For each individual, variant burden was quantified as the total number of non-reference alleles across these loci.

## Data Availability

All data produced in the present study are available upon reasonable request to the authors.

https://app.terra.bio/#workspaces/amp-pd-public/AMP-PD-In-Terra

## Acknowledgements

Human tissue was procured from the NIH NeuroBioBank at the following institutions: the University of Miami, the Mount Sinai Brain Bank, and the Harvard Brain Tissue Resource Center. Additional human tissue was obtained from the University of Miami Brain Endowment Bank and the University of Miami Udall Center of Excellence for Parkinson’s Disease Research. We are grateful to members of the Roussos laboratory for their insightful advice and critique, as well as to the Scientific Computing team at the Icahn School of Medicine at Mount Sinai for their computational resources and expertise.

This study was funded by grants from the National Institute of Neurological Disorders and Stroke (NINDS), including the National Institutes of Health (NIH) grant U01NS125580 (to P.R., V.H., and W.K.S.) and the Accelerating Medicine Partnership® (AMP®) Parkinson’s Disease (AMP PD) program (https://www.amp-pd.org).

## AMP PD Acknowledgement

Data used in the preparation of this article were obtained from the Accelerating Medicine Partnership® (AMP®) Parkinson’s Disease (AMP PD) Knowledge Platform. For up-to-date information on the study, visit https://www.amp-pd.org. The AMP® PD program is a public-private partnership managed by the Foundation for the National Institutes of Health and funded by the National Institute of Neurological Disorders and Stroke (NINDS) in partnership with the Aligning Science Across Parkinson’s (ASAP) initiative; Celgene Corporation, a subsidiary of Bristol-Myers Squibb Company; GlaxoSmithKline plc (GSK); The Michael J. Fox Foundation for Parkinson’s Research ; Pfizer Inc.; AbbVie Inc.; Sanofi US Services Inc.; and Verily Life Sciences.

ACCELERATING MEDICINES PARTNERSHIP and AMP are registered service marks of the U.S. Department of Health and Human Services.

## Author contributions

P.R. conceived and designed the study. D.A.D., S.B., V.H. and W.K.S., contributed to the selection of donors, obtained informed consents, reviewed clinical examinations and/or pathological analysis and dissected tissue. J.F.F. supervised data generation. T.C. designed downstream analysis. T.C. performed taxonomy annotations, differential expression and genetic correlation analysis, hotspot, TPS, cell-type compositional analysis, N.M. supported disease progression analysis. C.P. supported CCI analysis. T.C. and J.B supervised bioinformatics data analysis. P.N.M., J.B. and T.C. processed the raw sequencing data into the analysis-ready formats at the AMP PD Knowledge Platform. T.C., D.L., J.B. and P.R. wrote and edited the manuscript with input from all co-authors.

---

**Supplementary Fig. 1.**
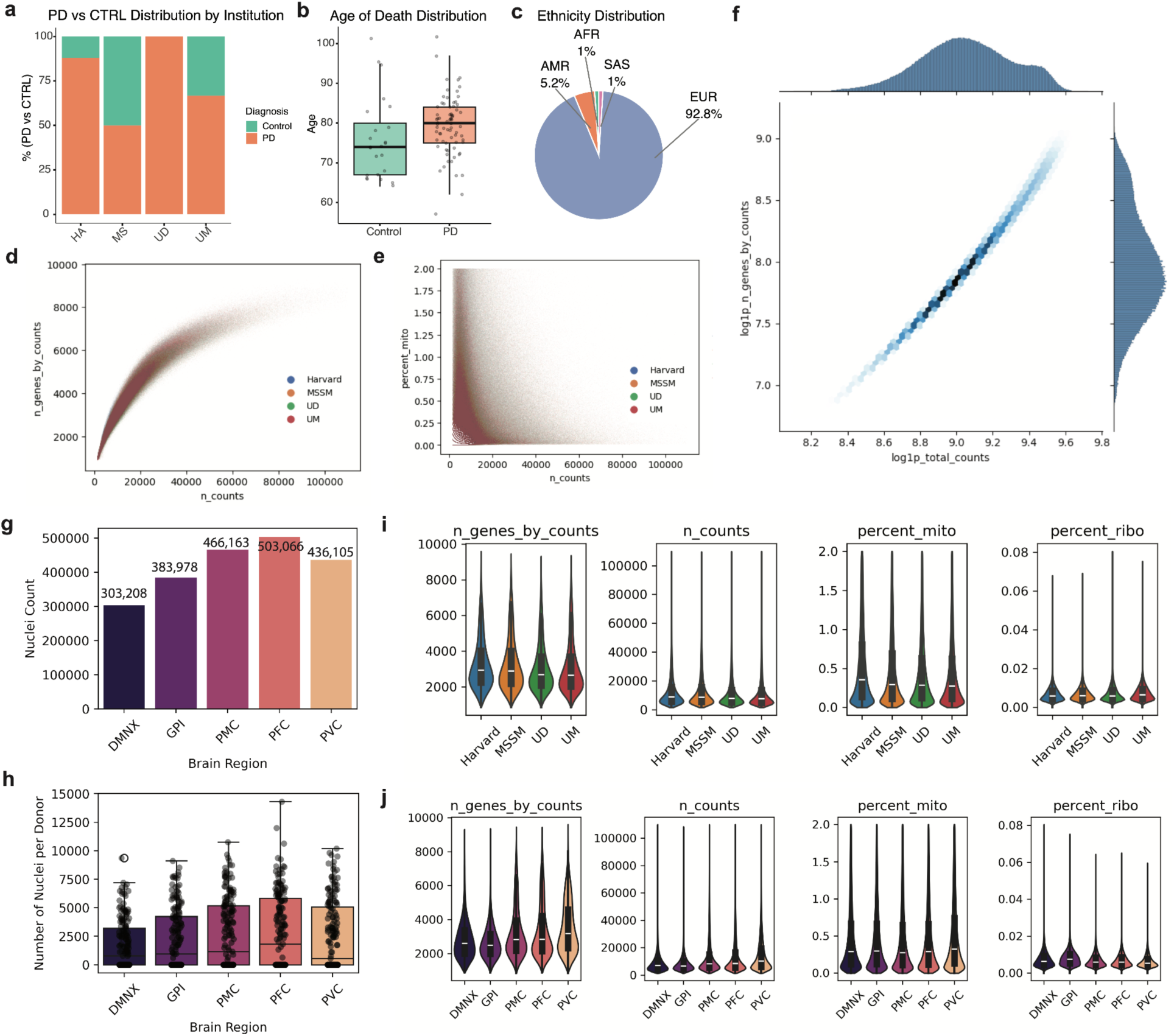
Institutional and biological heterogeneity, and single-nucleus quality metrics across the AMP-PD dataset. (a) *PD vs. control distribution across Institution sources;* Stacked bar plot showing the percentage of samples diagnosed with PD versus controls across contributing institutions (Harvard, MSSM, UD, UM). **(b)** *Age at death by diagnosis;* Comparison of age at death between PD and control donors. Boxes represent the interquartile range (IQR), the median is shown as a horizontal line, and whiskers extend to 1.5× IQR. Individual data points are overlaid. **(c)** *Ancestry distribution.* Proportion of donors by genetic ancestry: European (EUR), African (AFR), South Asian (SAS), and admixed American (AMR). Donors with unknown ancestry are not included. **(d)** *Gene count vs. UMI count per nucleus;* Relationship between the number of genes detected (n_genes_by_counts) and the total UMI count (n_counts) per nucleus, colored by institution. **(e)** *Mitochondrial content vs. UMI count;* Proportion of mitochondrial transcripts (percent_mito) as a function of total UMI counts, colored by institution. **(f)** *Total counts vs. gene counts (log-transformed);* Density-colored hex plot showing the correlation between log_10_-transformed gene and UMI counts across all nuclei, with marginal density plots. **(g)** *Nuclei count per brain region.* Total nuclei captured from each sampled brain region: dorsal motor nucleus of the vagus (DMNX), globus pallidus interna (GPI), primary motor cortex (PMC), prefrontal cortex (PFC), and primary visual cortex (PVC). **(h)** *Nuclei per donor across brain regions;* Distribution of nuclei per donor across each brain region. Boxes represent IQR, and whiskers extend to 1.5× IQR. **(i)** *QC metrics across institutions;* Distribution of four quality control metrics per institution: number of genes (n_genes_by_counts), total UMI counts (n_counts), mitochondrial gene percentage (percent_mito), and ribosomal gene percentage (percent_ribo). **(j)** *QC metrics across brain regions;* Violin plots showing the same four QC metrics as in (i), stratified by brain region.

**Supplementary Fig. 2.**
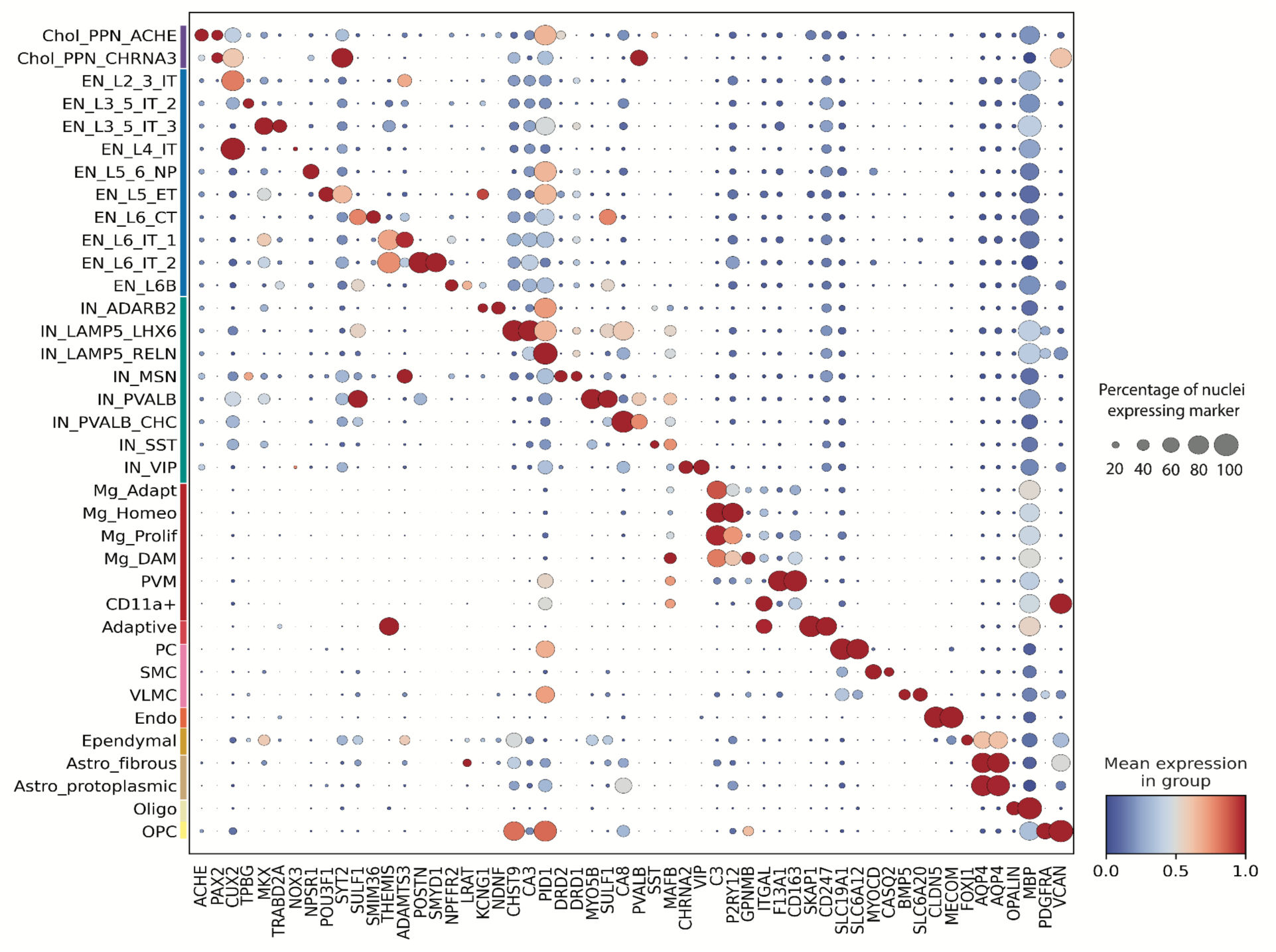
Representative subclass markers of AMP-PD dataset. Dot plot visualizing representative marker genes for each subclass in the taxonomy. The size of each dot indicates the percentage of nuclei expressing the gene, while the color intensity reflects the mean expression in the relevant group. This representation highlights subclass-specific expression patterns across the transcriptomic hierarchy.

**Supplementary Fig. 3.**
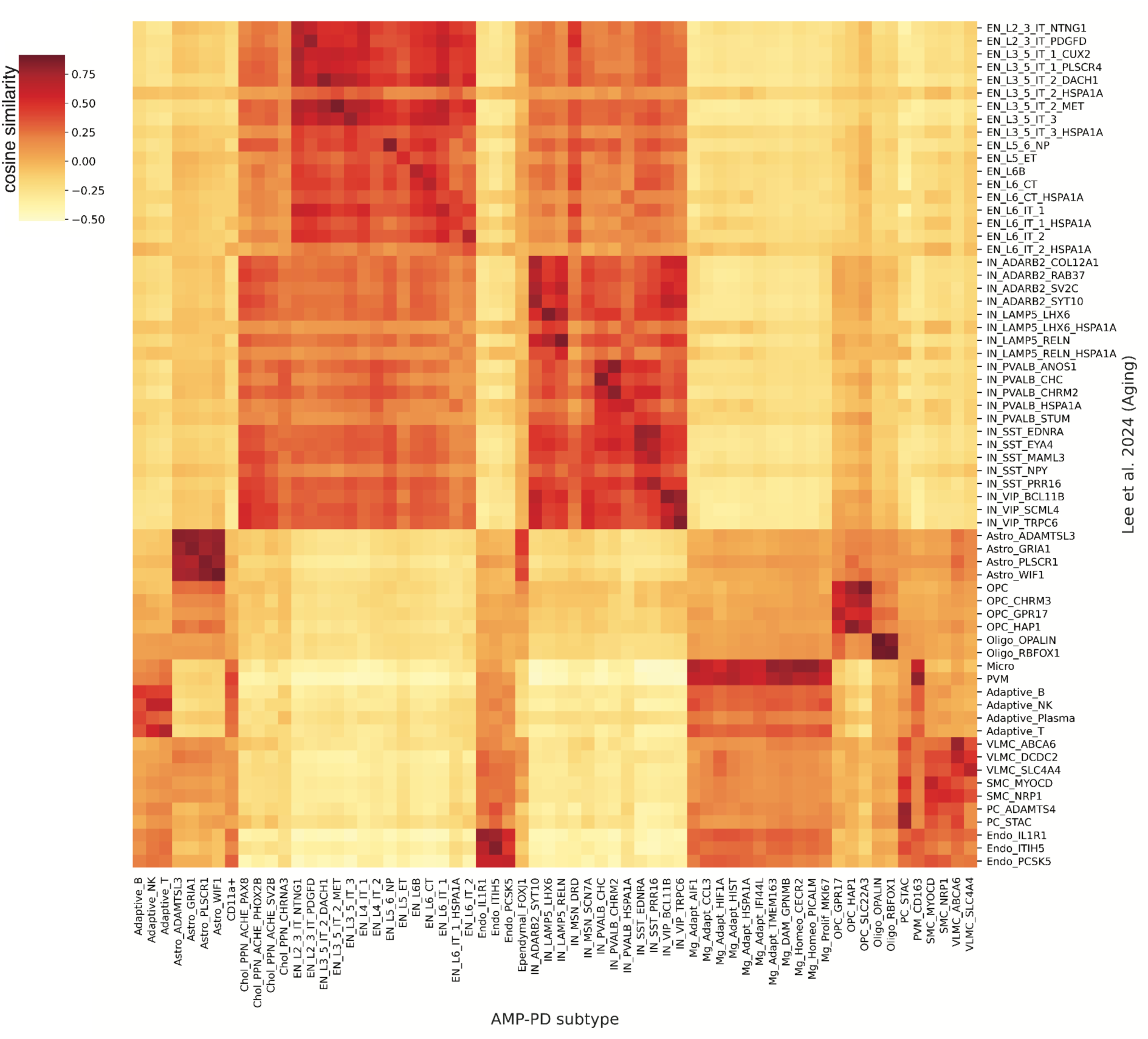
Cross-cohort subtype similarity analysis. Cosine similarity matrix comparing pseudobulk transcriptomic profiles at the subtype level between the AMP-PD cohort and the Aging cohort from Lee et al. (2024)^20^. This analysis quantifies the degree of transcriptional correspondence between subtypes across datasets.

**Supplementary Fig. 4.**
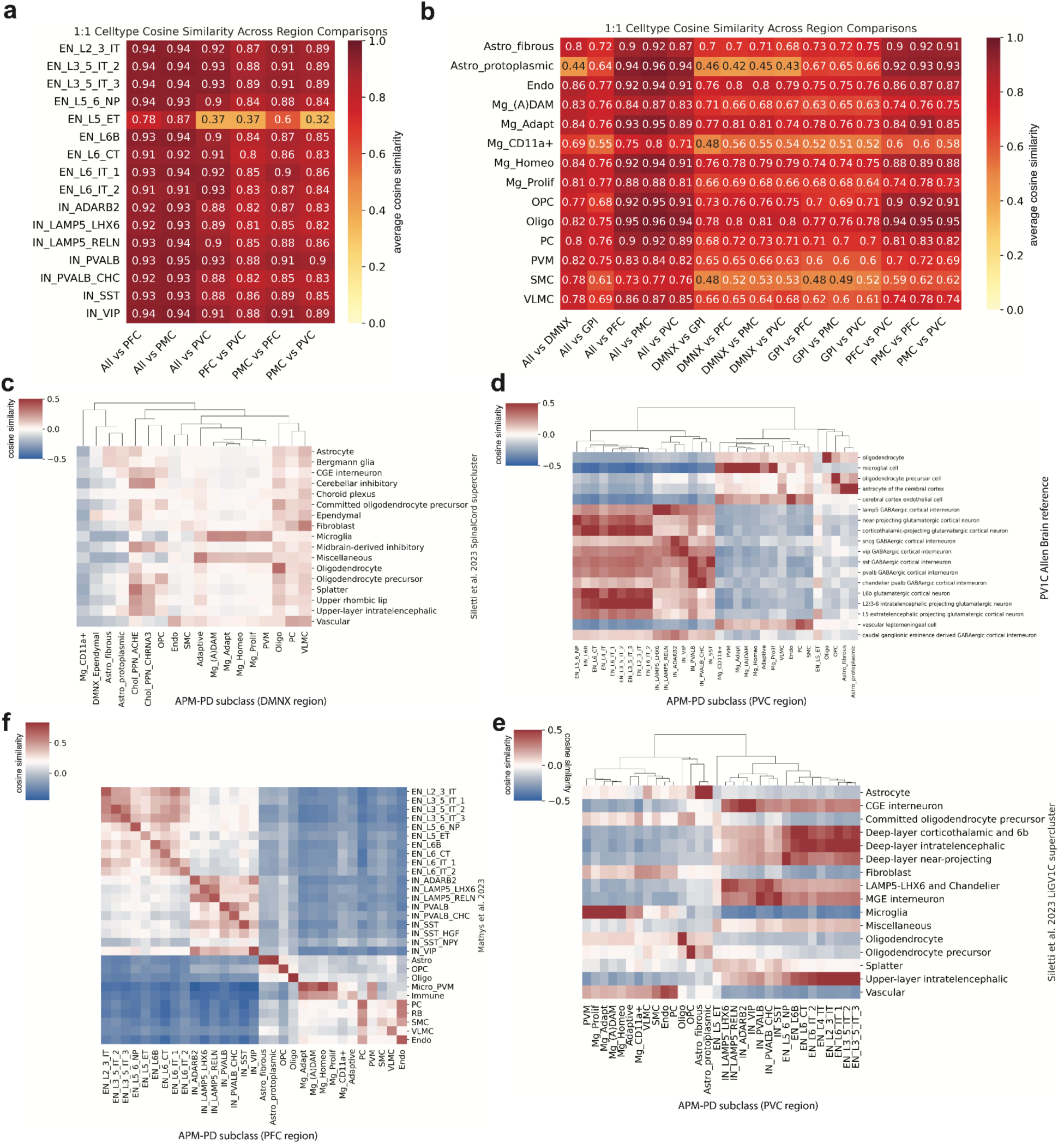
(a) *Cosine similarity of neuronal subclasses across brain regions.* Heatmap showing pairwise cosine similarity between pseudobulk profiles of neuronal subclasses across regions. Excitatory and inhibitory neuronal populations are shown, highlighting region-specific variation in transcriptional profiles among neuronal types. **(b)** *Cosine similarity of glial subclasses across brain regions.* Heatmap showing pairwise cosine similarity between average pseudobulk expression profiles of shared glial cell subclasses across brain regions. Subclasses include astrocytes, oligodendrocytes, microglia, and other non-neuronal populations. Higher similarity (red) indicates more conserved transcriptional identity across anatomical regions. **(c-f)** *Transcriptomic similarity between AMP-PD subclasses and external reference datasets.* Cosine similarity heatmaps comparing AMP-PD subclass pseudobulk profiles from specific regions to reference datasets: **(c)** DMNX region vs. spinal cord region (ScP) from Siletti et al. (2023)^26^, **(d)** PVC region vs. PV1C Allen Brain dataset^27^, **(e)** PFC region vs. Alzheimer’s dataset from Mathys et al. (2023)^21^, **(f)** PVC region vs. LiGV1C region from Siletti et al. (2023)^26^. Rows indicate AMP-PD subclasses; columns indicate matched or analogous cell types in the reference datasets.

**Supplementary Fig. 5.**
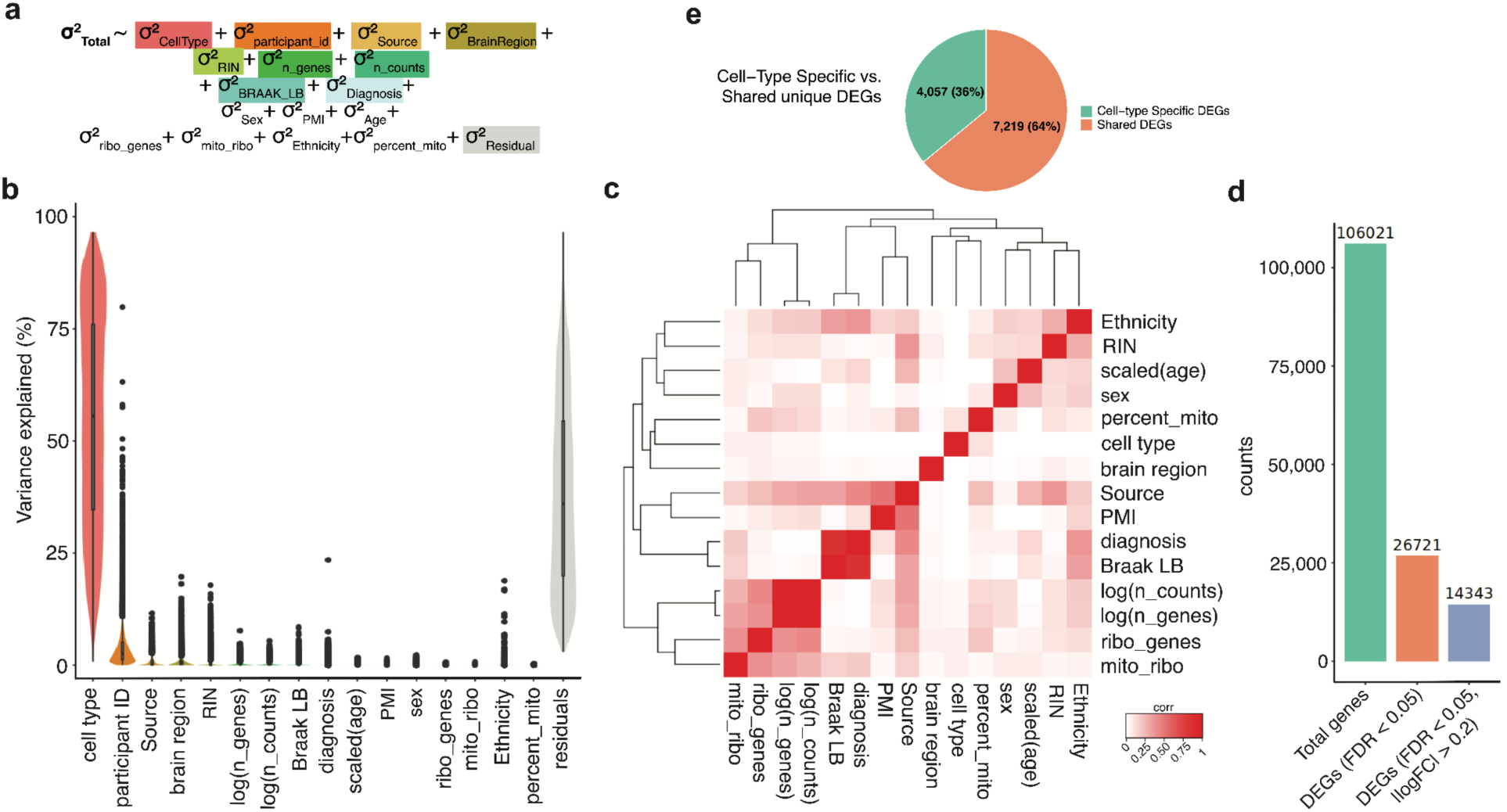
Partitioning of Gene Expression Variance and Cell Type-Specific DEG Overlap in PD. (a) *Equation summarizing the total contribution of covariates to gene expression variance*, highlighting the individual effects of biological and technical factors. PMI: Post-mortem interval, RIN: RNA integrity number. (b) *Gene expression variance*, incorporating technical and donor-level covariates, as described in Hoffman and Schadt et al. (2016)^28^. (c) *Pairwise correlation matrix of donor-level metadata*, highlighting relationships among them. (d) *Number of DEGs under filtering criteria* (total in green, FDR <0.05 in orange and FDR < 0.05 and|log_2_FC|>0.2 in blue). (e) *Overlap of significant DEGs (FDR < 0.05 and |log_2_FC| > 0.2) across cell types*, distinguishing cell type-unique DEGs from shared DEGs.

**Supplementary Fig. 6.**
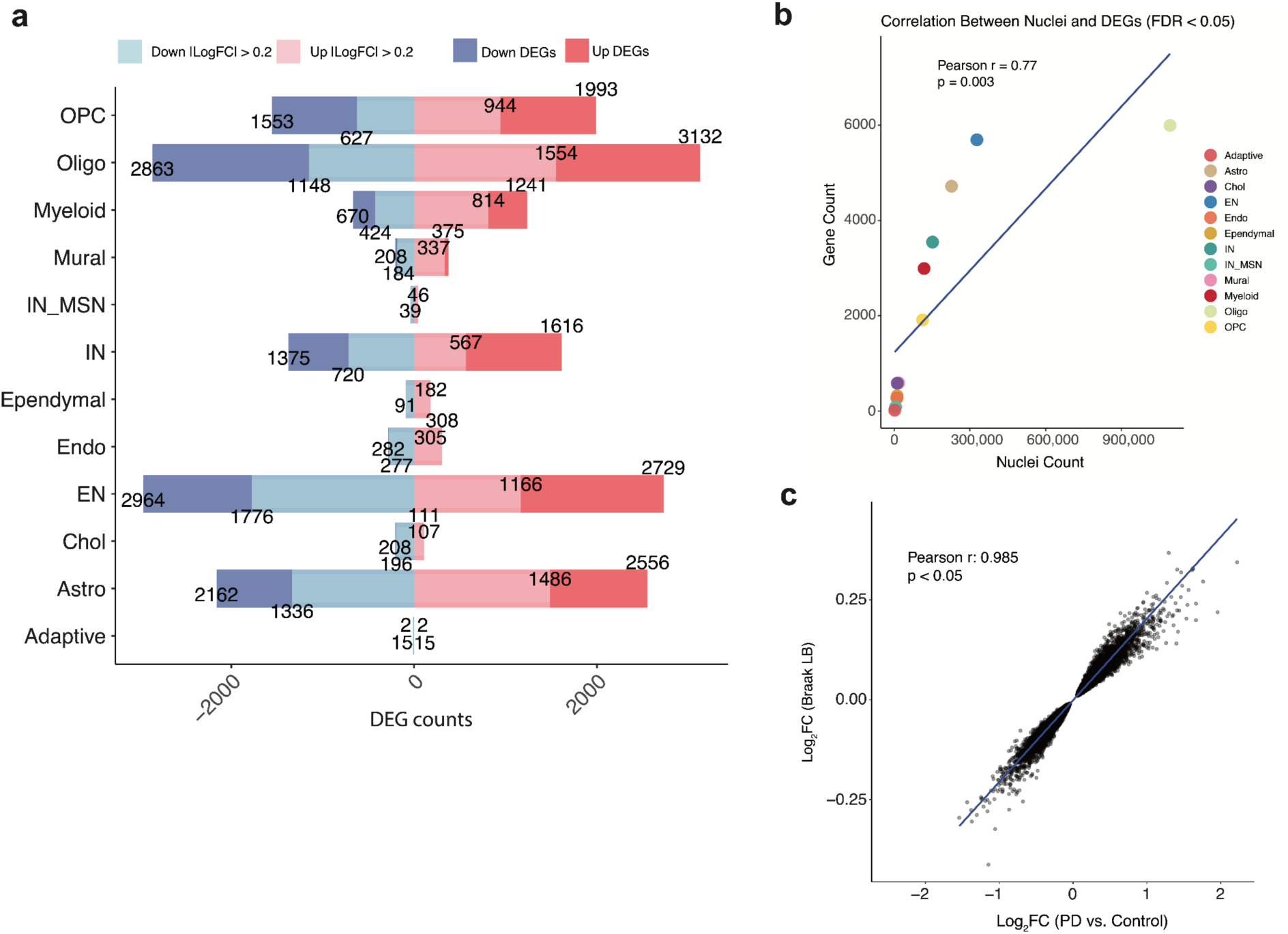
(a) *Number of DEGs per cell type*, with significant DEGs defined by FDR < 0.05 and |log_2_FC| > 0.2. Red bars indicate up-regulated DEGs, while blue bars indicate down-regulated DEGs. (b) *Concordance* between log₂FC of significant DEGs (FDR < 0.05) in PD vs. Control and Braak LB DE analyses (Pearson *r* = 0.99, p < 0.05). (c) *Correlation* between nuclei count per cell class and the number of significant DEGs (FDR < 0.05) in PD vs. Control (Pearson *r* = 0.77, *p* = 0.003).

**Supplementary Fig. 7.**
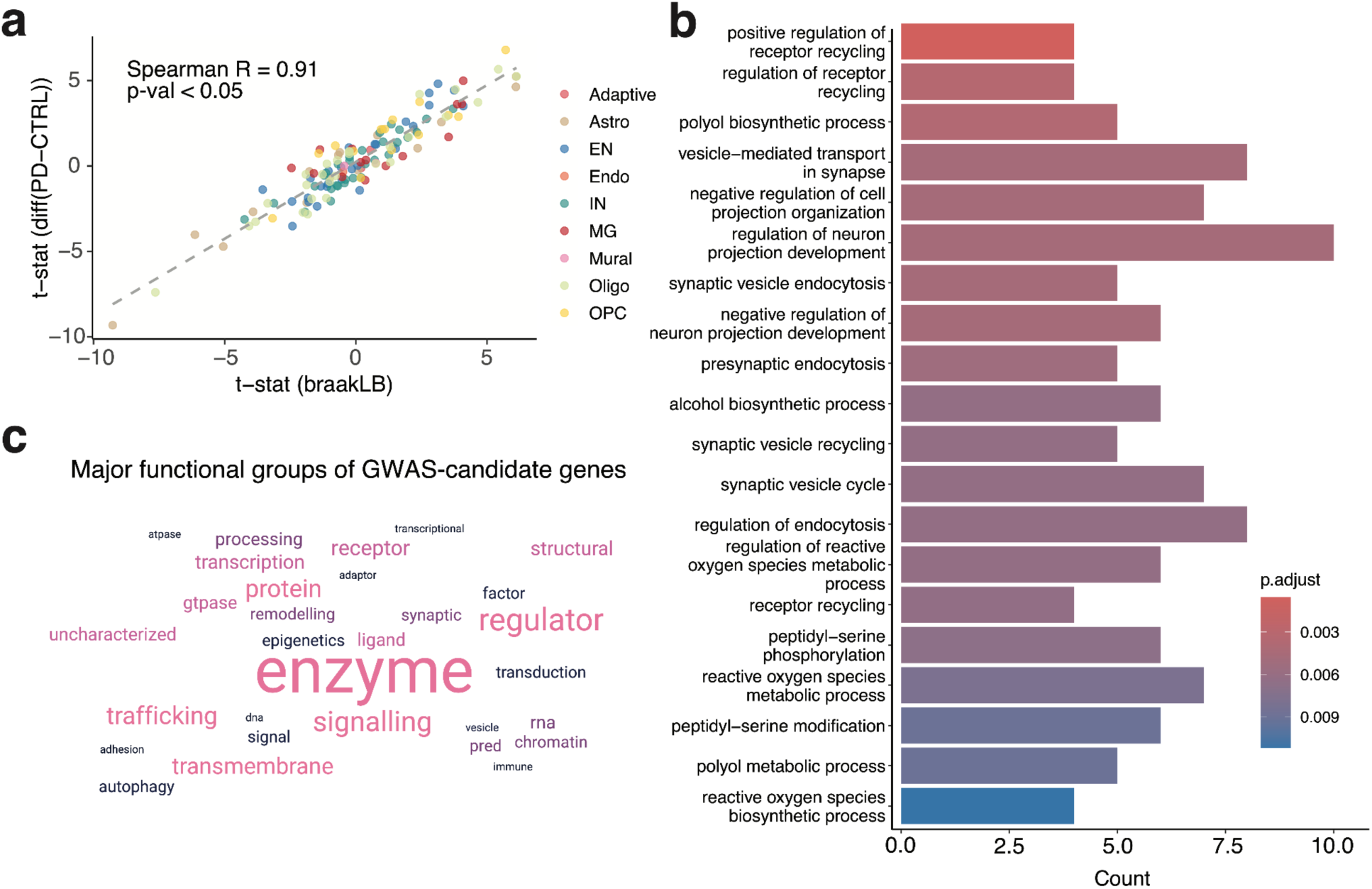
PD-GWAS transcriptomic integration and functional interpretation across brain cell types. (a) *DE t-statistics* from PD vs control and Braak LB progression comparisons across PD-GWAS genes (ρ = 0.91, *P* < 0.05). (b) *GO enrichment of biological processes* for the 84 PD-GWAS candidates identifies vesicle cycling, endocytosis, and neuronal morphogenesis pathways. (c) *Functional annotation* word cloud of 84 PD-GWAS candidates.

**Supplementary Fig. 8.**
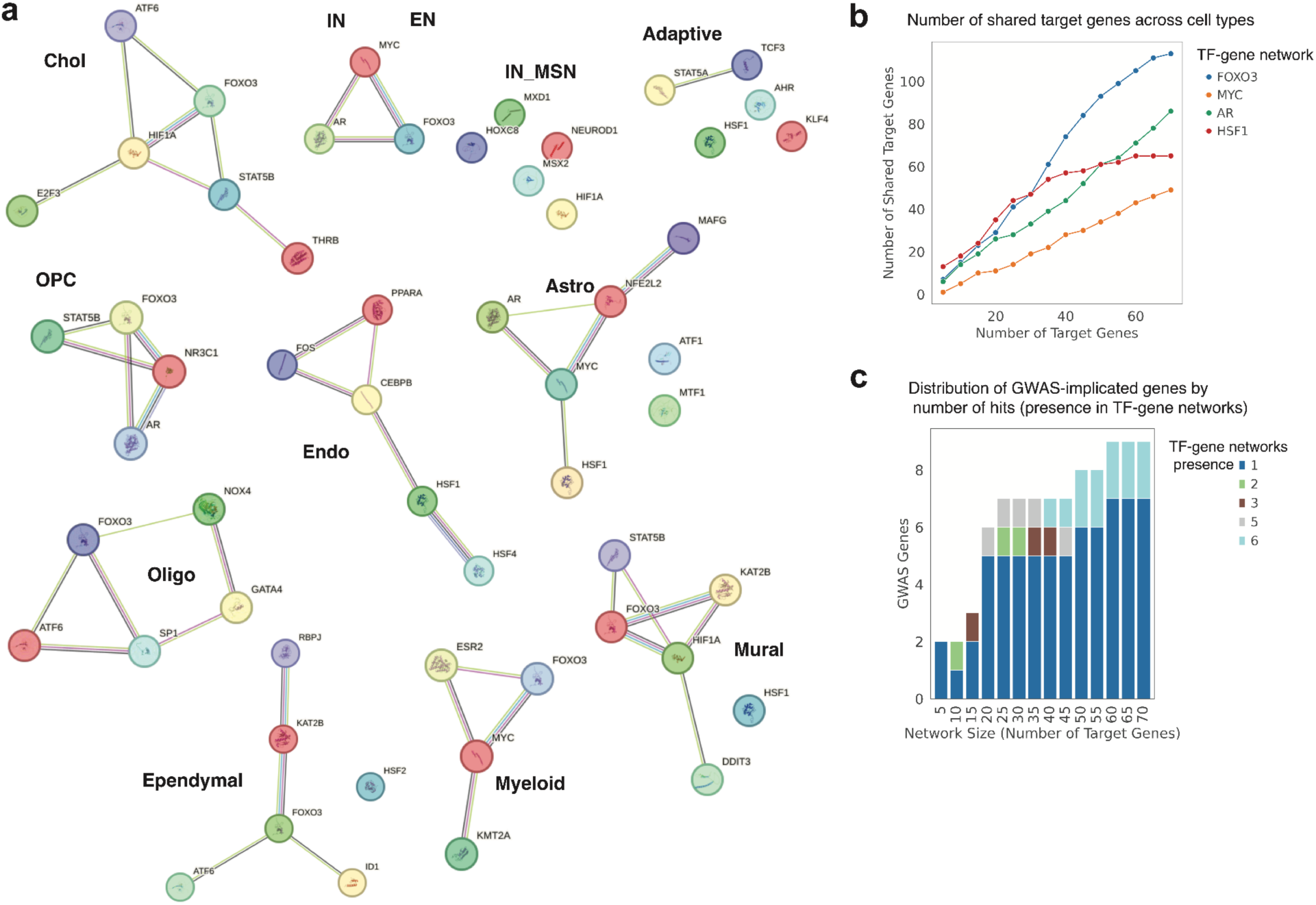
Shared transcriptional regulators and gene targets across cell types. (a) *STRING protein-protein interaction* (PPI) network^50^ visualization of top up-regulated TFs per cell type. Colored nodes represent query TFs and their first shell of interactors. Nodes representing TFs with known or predicted 3D structures are shown with filled circles; distinct colors indicate structurally distinct TFs. STRING network edges indicate different types of functional associations, with curated interactions from databases (light blue) and experimental evidence (pink), while computationally inferred associations include gene neighborhood (green), gene fusions (red), gene co-occurrence (blue), text mining (yellow), co-expression (black), and protein homology (purple). **(b)** *Number of shared target genes* across cell types for key multi-lineage TFs (HSF1, MYC, AR, FOXO3) as a function of TF-target network size. **(c)** *Number of GWAS-implicated genes found in TF-target gene networks* across increasing network sizes, stratified by the number of distinct TF-cell type networks in which they appear. Most genes were uniquely assigned to a single regulatory context, reinforcing specificity.

**Supplementary Fig. 9.**
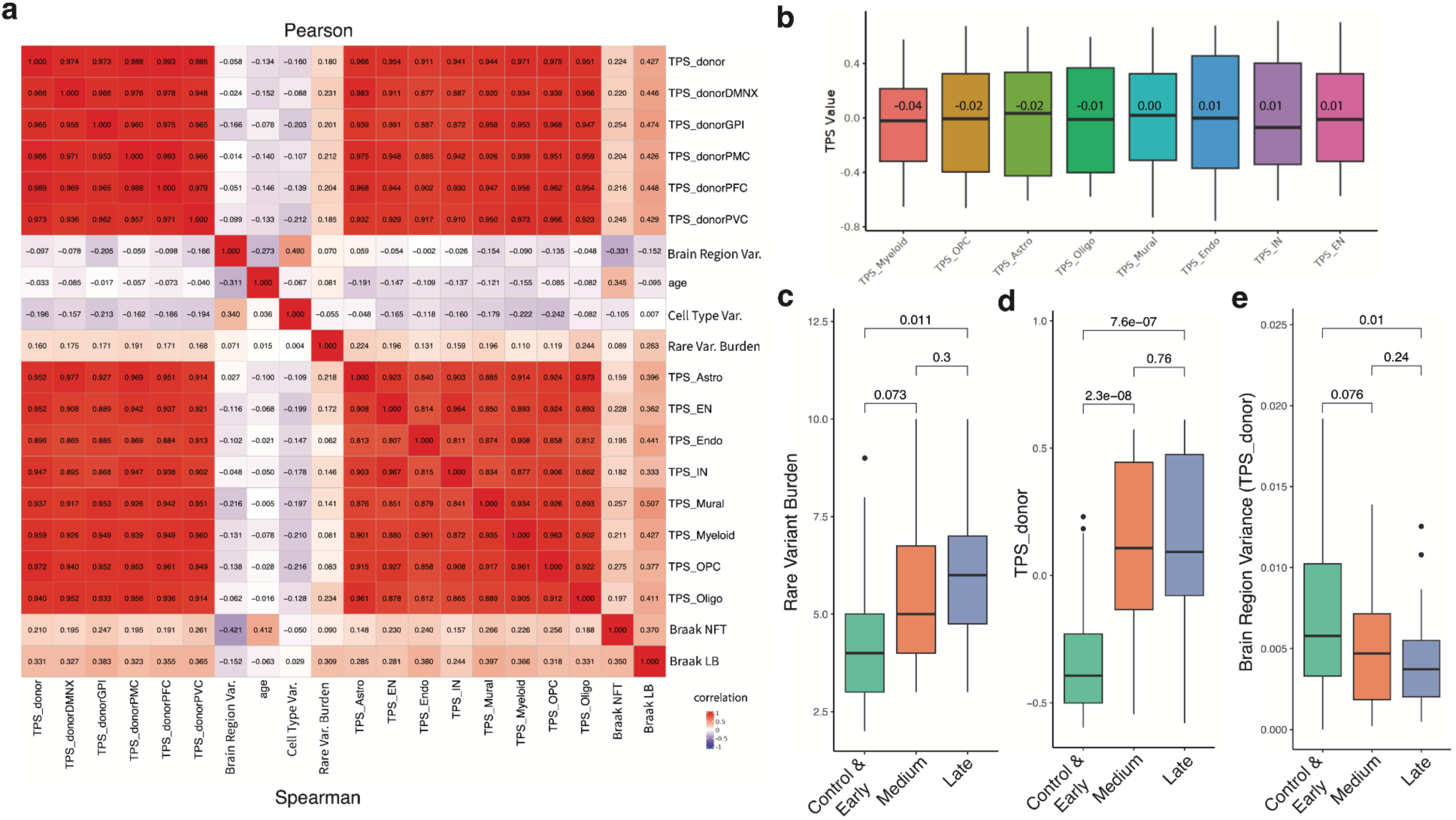
Correlation and distribution of Transcriptomic Pathology Score (TPS) across brain regions and cell types. (a) *Correlation heatmap* showing relationship between various TSP scores (cell type and brain-region specific) and selected clinical metadata using Spearman (lower triangle) and Pearson (upper triangle) correlation. **(b)** *Distribution of TPS values by cell type*, ordered by their mean TPS. **(c-e)** Rare variant burden (c), donor-level TPS (d) and brain region variance in TPS (e) across Braak LB stages. Statistical significance was assessed using pairwise t-tests, with p-values indicated for each comparison.

**Supplementary Fig. 10.**
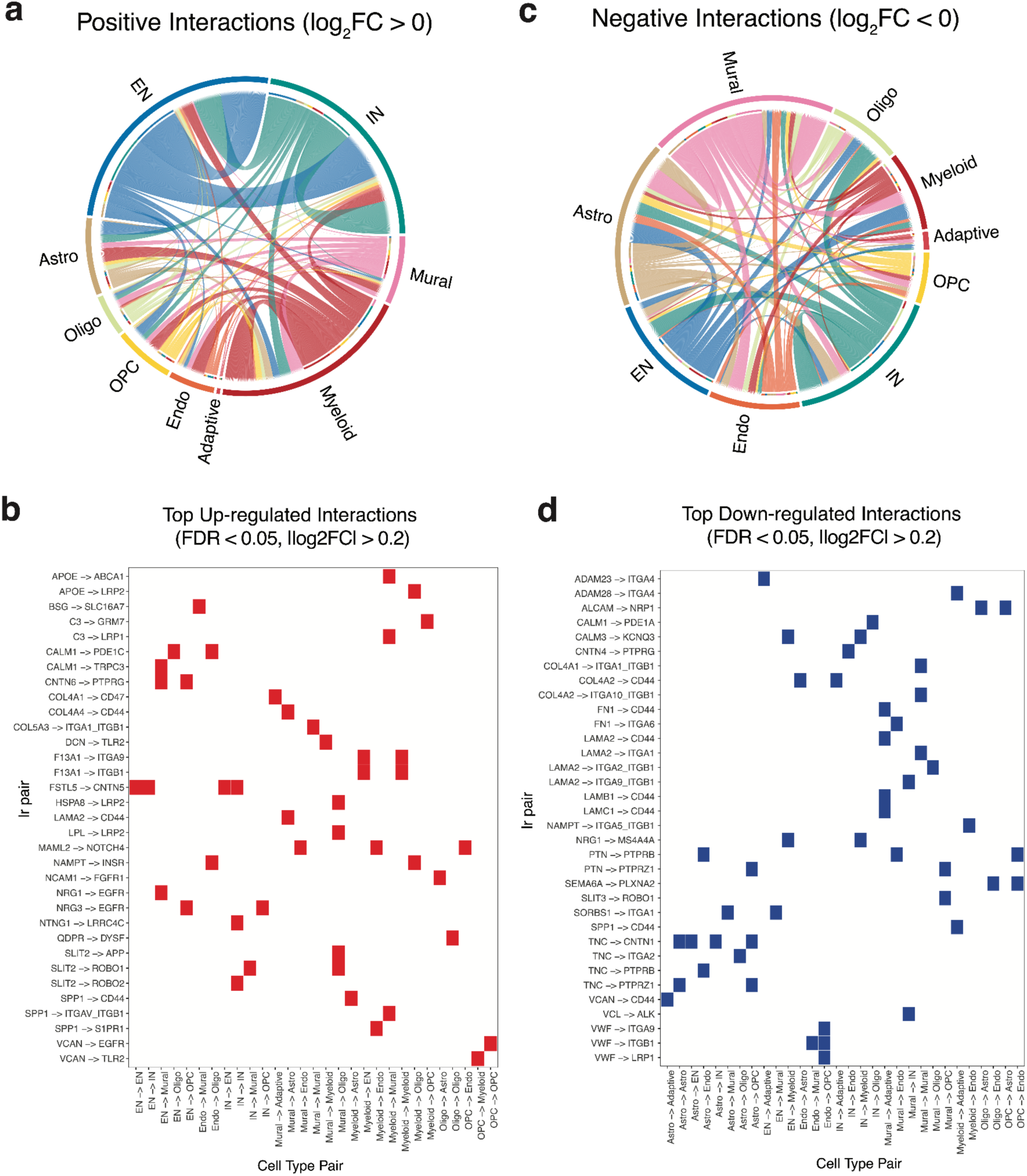
Visualization of positive and negative intercellular signaling programs and top dysregulated LR interactions. (a,c) *Chord diagrams summarizing positive (log₂FC > 0) and negative (log₂FC < 0) CCIs* across all cell types, showing increased signaling to and from neuronal and myeloid populations in PD. Connection colors denote the sender cell type, providing directional context to the observed interactions. **(b,d)** *Heatmaps of the most significantly up-(c) and down-regulated (d) LR pairs* (FDR < 0.05, |log₂FC| > 0.2), annotated by ligand–receptor identity and sender–receiver cell type pairs.

**Supplementary Fig. 11.**
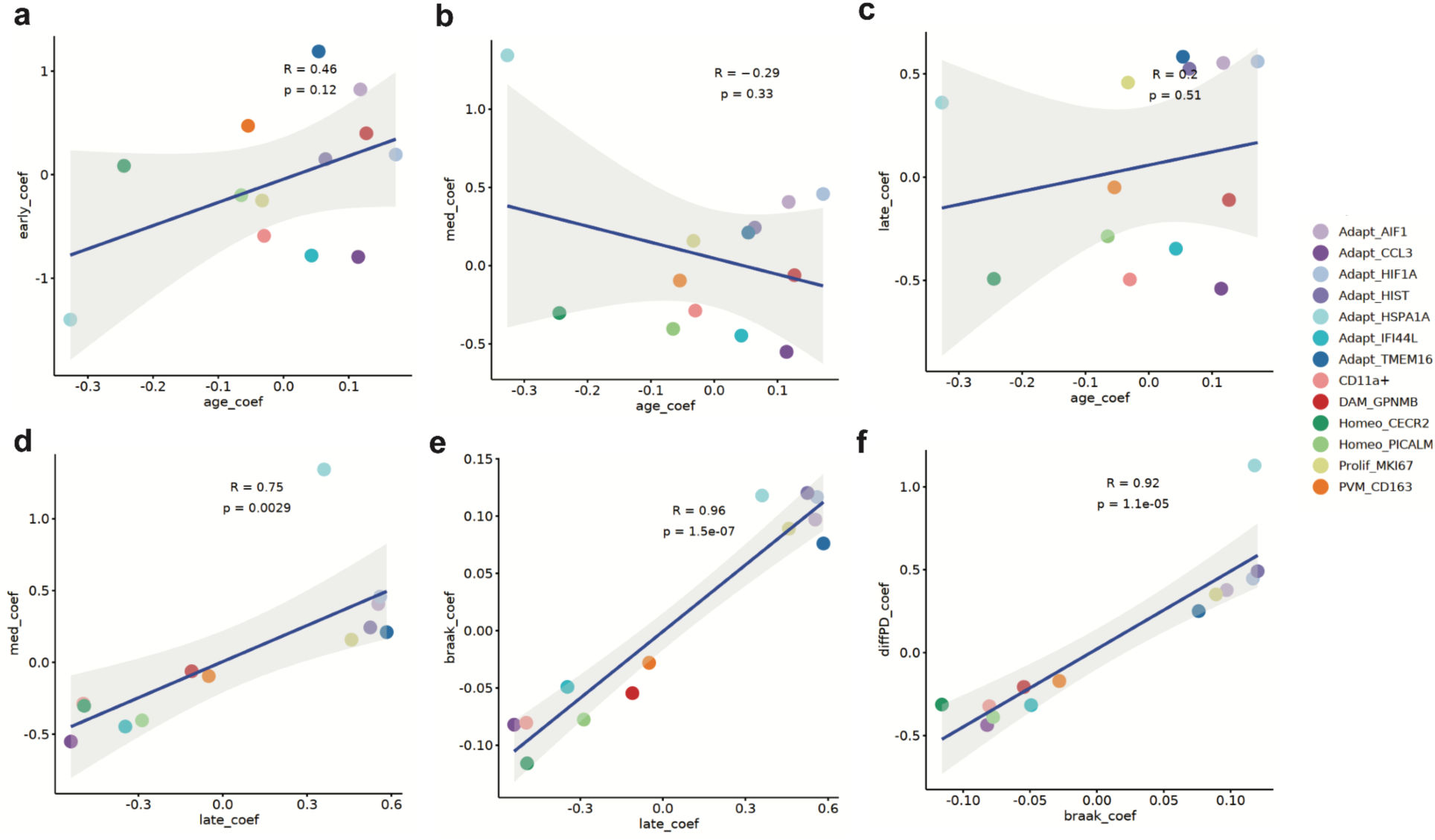
**(a-c)** *Comparison of subtype-level compositional changes* across early, medium, and late Braak LB stages to age-associated changes, with Pearson correlations of *r* = 0.46, -0.29, and 0.20 (*p* = 0.12, 0.33, and 0.51), respectively. **(d-f)** *Additional comparisons show stronger concordance of subtype shifts* between medium and late Braak LB stages (*r* = 0.75, *p* = 0.0029), late-stage Braak LB and Braak LB progression pseudotime (*r* = 0.96, *p* = 1.5 × 10⁻⁷), and PD vs. control compared with Braak LB progression (*r* = 0.92, *p* = 1.1 × 10⁻⁵), highlighting the trajectory of myeloid compositional remodeling across disease progression.

**Supplementary Fig. 12.**
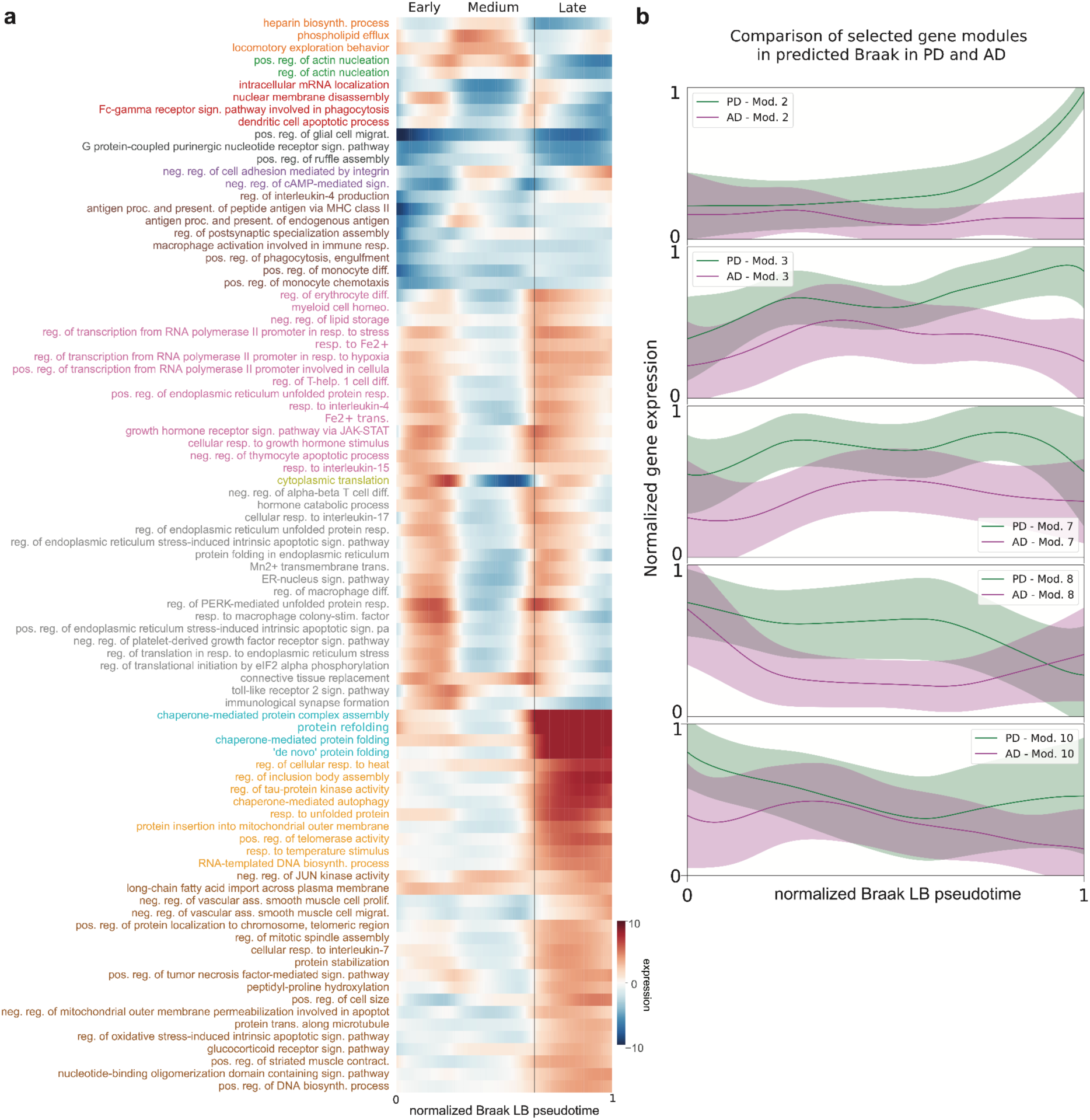
(a) *Change in pathway gene expression calculated using a sliding window across predicted Braak LB pseudotime*. Hue indicates Zenith z-score. Font color indicates pathway cluster membership. The selected list of pathways is shown in **Extended Data** Fig. 7e. **(b)** *Comparative analysis of selected gene co-expression modules across predicted Braak pseudotime* in PD (green) and AD (purple). Smoothed line plots show the normalized module expression dynamics across predicted disease progression. Modules were selected based on enrichment for key biological processes and are matched between diseases by module number as shown in Fig. 7f**-g**.

**Extended Data Figure 1.**
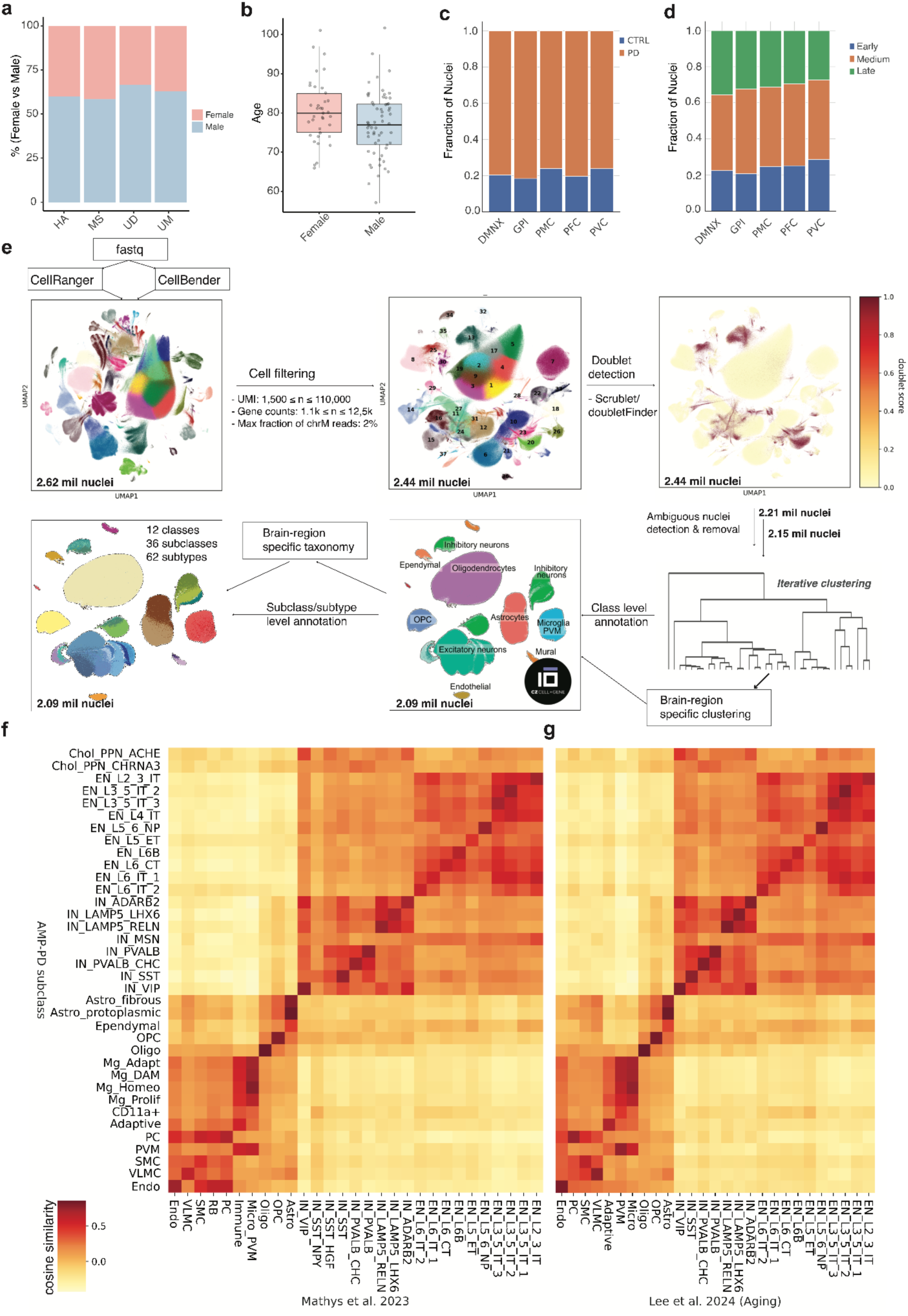
Cohort composition, regional sampling, and preprocessing workflow for the AMP-PD dataset. (a) *Sex representation by brain bank;* Male-to-female ratio across different brain banks/institutional sources contributing to the dataset. **(b)** *Age at death by sex;* Distribution of donor age at death stratified by sex. Each box represents the interquartile range (IQR), with the median indicated by a horizontal line. Whiskers extend to 1.5× IQR, and outliers are displayed as individual points. **(c)** *Regional representation of disease status;* Fraction bar plot showing the proportion of PD and control samples across five brain regions. **(d)** *Regional distribution of LB pathology severity;* Fractions of samples from each Braak LB group-early, medium, and late - across the same brain regions. **(e)** *snRNA-seq preprocessing and annotation overview;* Stepwise schematic illustrating data preprocessing, quality control, hierarchical clustering, and cell type taxonomy generation for the AMP-PD atlas. Each window represents a major step in the computational pipeline. **(f-g)** *Cross-cohort subclass-level transcriptomic similarity;* Cosine similarity heatmaps comparing pseudobulk transcriptomic profiles at the subclass level between the AMP-PD dataset and two external snRNA-seq datasets: the AD cohort from Mathys et al. (2023)^21^ (f) the Aging (control) cohort from Lee et al. (2024)^20^ (g).

**Extended Data Figure 2.**
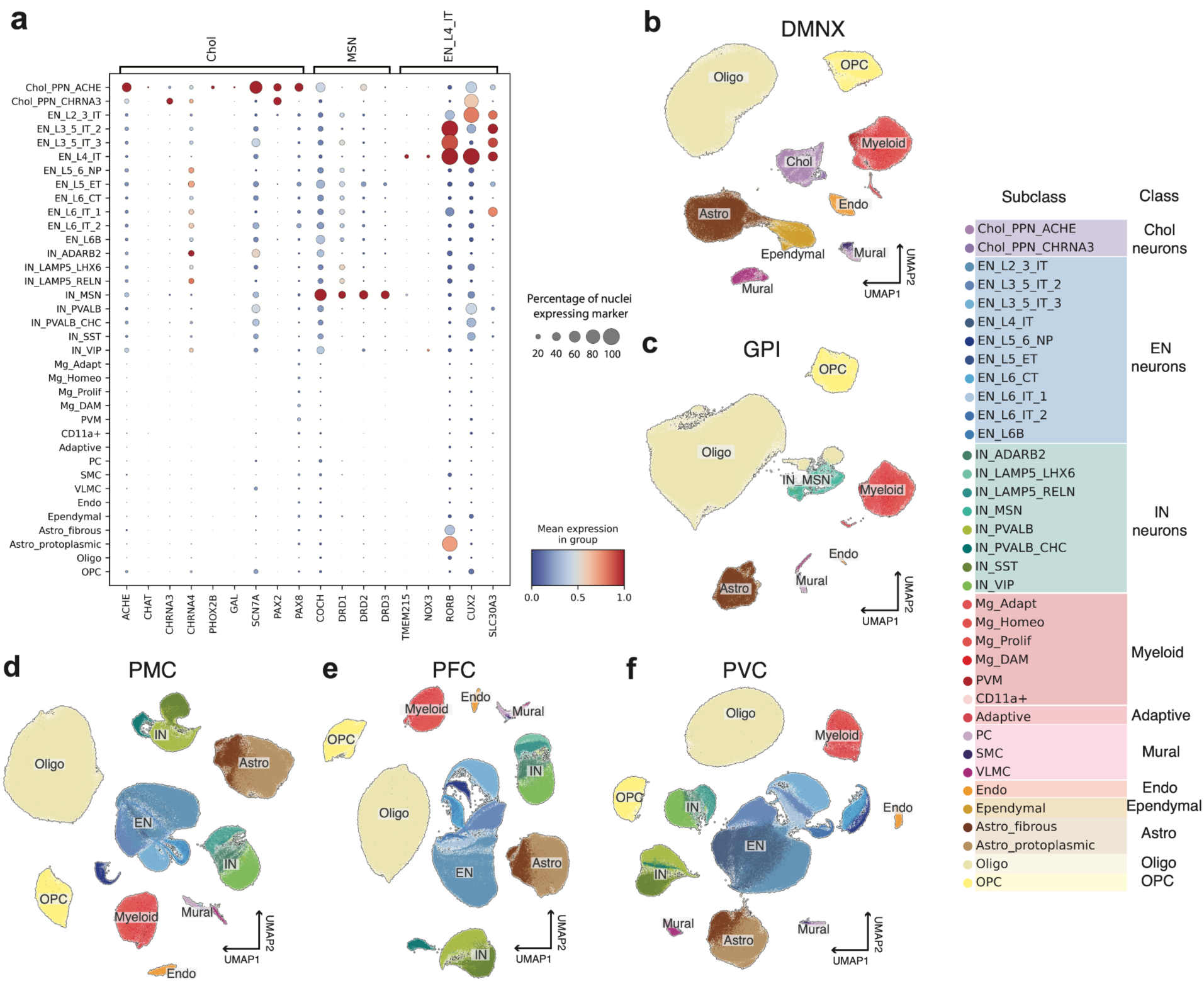
Regional resolution of subclass-level taxonomy across brain regions. (a) *Marker expression of region-specific neuronal subclasses.* Dot plot displaying expression of marker genes specific to cholinergic (Chol), medium spiny neurons (MSN), and excitatory L4 intratelencephalic (EN_L4_IT) neuron populations. Dot size represents the fraction of nuclei expressing each gene, while color indicates the average z-score normalized expression within each subclass-level cell type. **(b-f)** *UMAP embeddings of subclass-level taxonomy per brain region -* **(b)** DMNX, **(c)** GPI, **(d)** PMC, **(e)** PFC, and **(f)** PVC. Each UMAP shows nuclei colored by subclass identity as defined in the unified taxonomy.

**Extended Data Figure 3.**
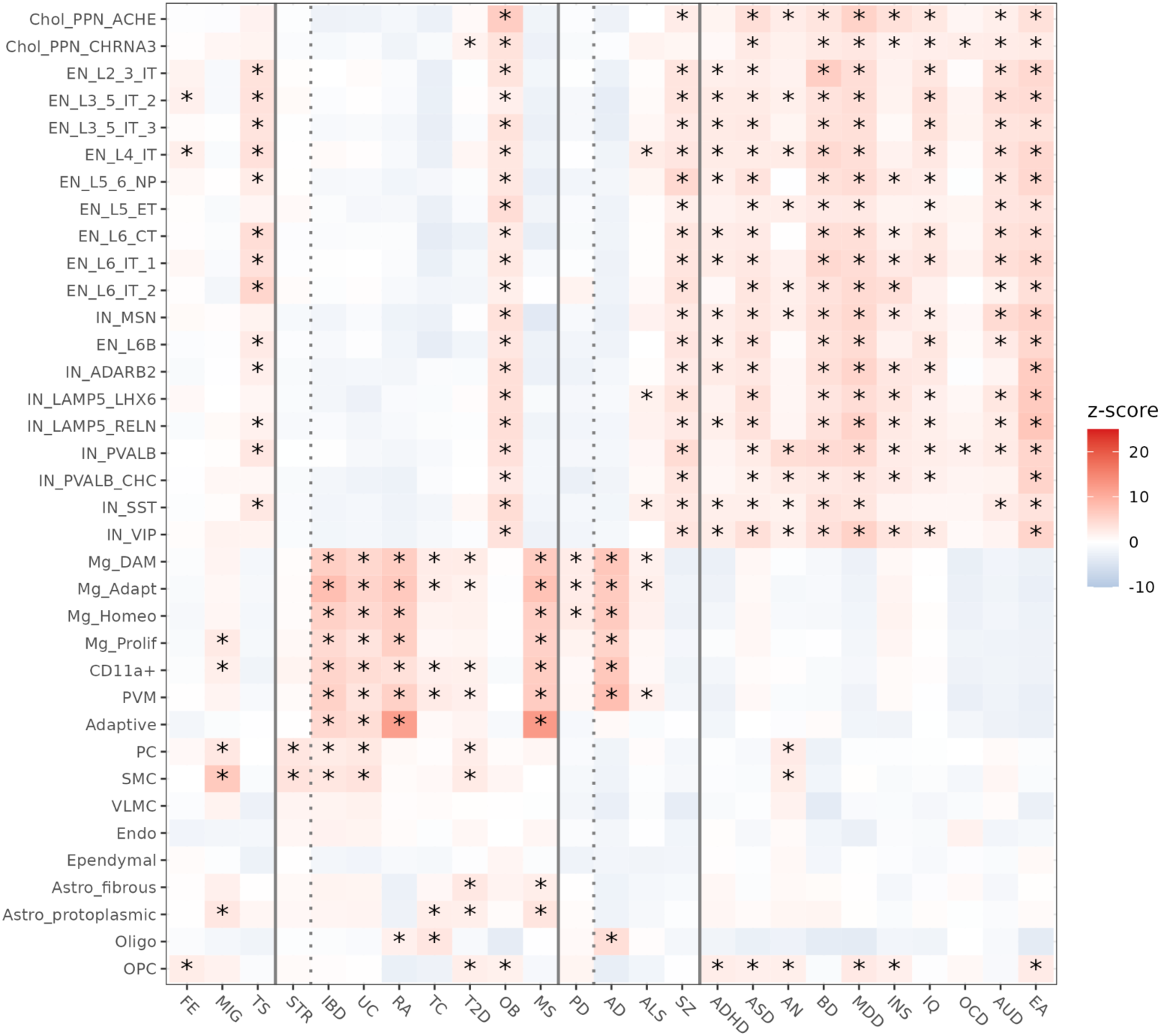
Subclass-level scDRS associations across neurological, metabolic and immune, neurodegenerative and psychiatric traits. Heatmap showing scDRS z-scores for subclass-specific associations across 25 GWAS traits (**Supplementary Table 3**). Rows represent neuronal, glial, vascular, and immune subclasses; columns correspond to traits. Color scale indicates the magnitude of z-scores (blue, negative; red, positive). Asterisks (*) denote statistical significance after multiple testing corrections (FDR < 0.05). Vertical lines separate trait domains - neurological, metabolic and immune (and neurodegenerative), psychiatric traits. Trait abbreviations: FE (focal epilepsy), MIG (migraine), TS (Tourette syndrome), STR (stroke), IBD (inflammatory bowel disease), UC (ulcerative colitis), RA (rheumatoid arthritis), TC (total cholesterol), T2D (type 2 diabetes), OB (obesity), MS (multiple sclerosis), PD (Parkinson’s disease), AD (Alzheimer’s disease), ALS (amyotrophic lateral sclerosis), SZ (schizophrenia), ADHD (attention-deficit/hyperactivity disorder), ASD (autism spectrum disorder), AN (anorexia nervosa), BD (bipolar disorder), MDD (major depressive disorder), INS (insomnia), IQ (intelligence), OCD (obsessive-compulsive disorder), AUD (alcohol use disorder), EA (educational attainment).

**Extended Data Figure 4.**
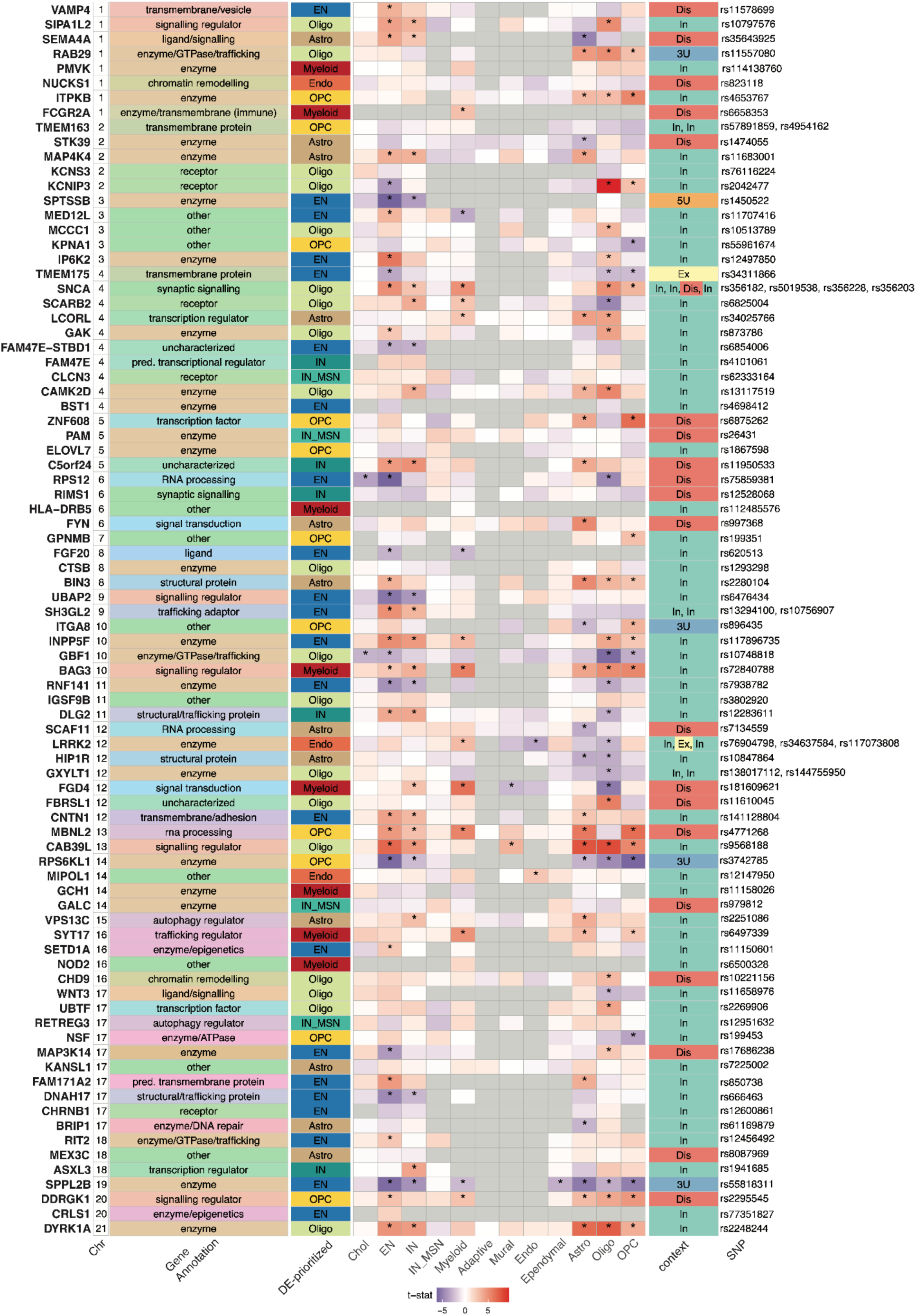
Genetic, functional and transcriptional context of PD-GWAS candidate genes across brain cell types. Heatmap summarizing 84 FDR-significant candidate genes from PD-GWAS that were also detected in single-nucleus differential expression (DE) analysis. Genes are arranged by genomic position (chromosomes 1–22). Annotations include overlap with PD-TWAS (green = present, white = not detected), simplified functional categorization, and gene-specific DE t-statistics across major class level brain cell types. Asterisks (*) denote FDR-significant differential expression (*FDR* < 0.05). For each gene, the cell type with the largest absolute t-statistic is indicated. The final columns provide genetic context, including locus-level classification of the associated GWAS signal (exonic [Ex], intronic [In], distal intergenic [Dis], 3′ UTR [3U], or 5′ UTR [5U]) and the lead variant at the risk locus.

**Extended Data Figure 5.**
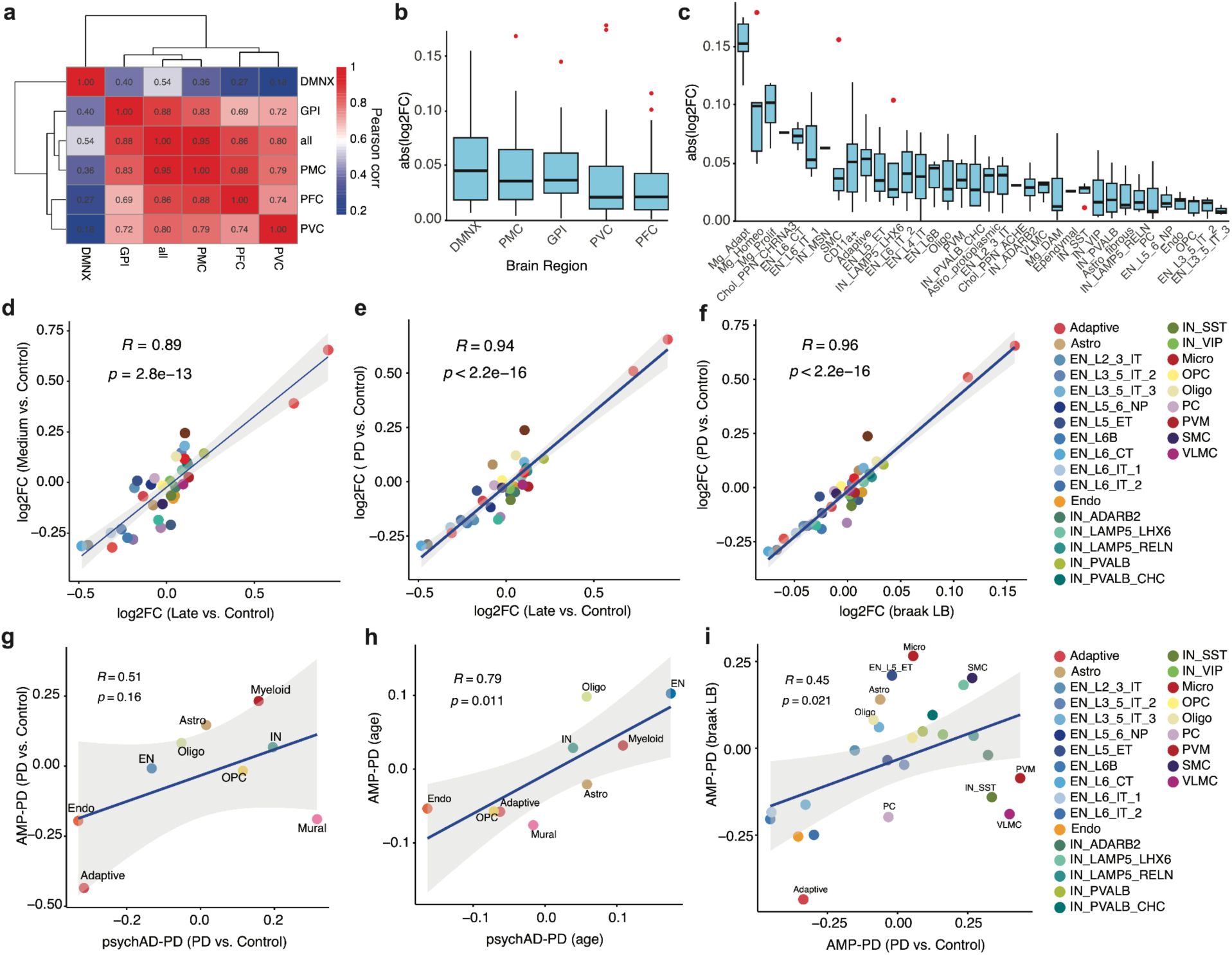
Cell-type compositional analysis across brain regions and validation in an independent replication cohort. (a) Correlation heatmap of PD vs Control effect sizes across different brain regions, highlighting shared and distinct regional trends. (b) PD vs Control log_2_FC, ordered by the median effect size across brain regions. (c) Distribution of PD vs. Control effect sizes by subclass, showing variability in subclass-specific responses across regions. **(d-f)** Comparison of effect sizes across various conditions; Medium vs Control (d), PD vs Control versus Late vs Control (e), and PD vs Control versus Braak LB (f) from composition analysis using crumblr ^24^. **(g-h)** Validation of cell-type composition changes in an independent replication psychAD cohort (Lee et al. 2024^20^) using crumblr analysis, for PD vs Control contrast (g) and scaled aged (h). **(i)** Comparison of crumblr-estimated PD effect sizes with Braak LB log_2_FC, highlighting concordance between clinical diagnostic status and neuropathology-based analyses within the AMP-PD dataset.

**Extended Data Figure 6.**
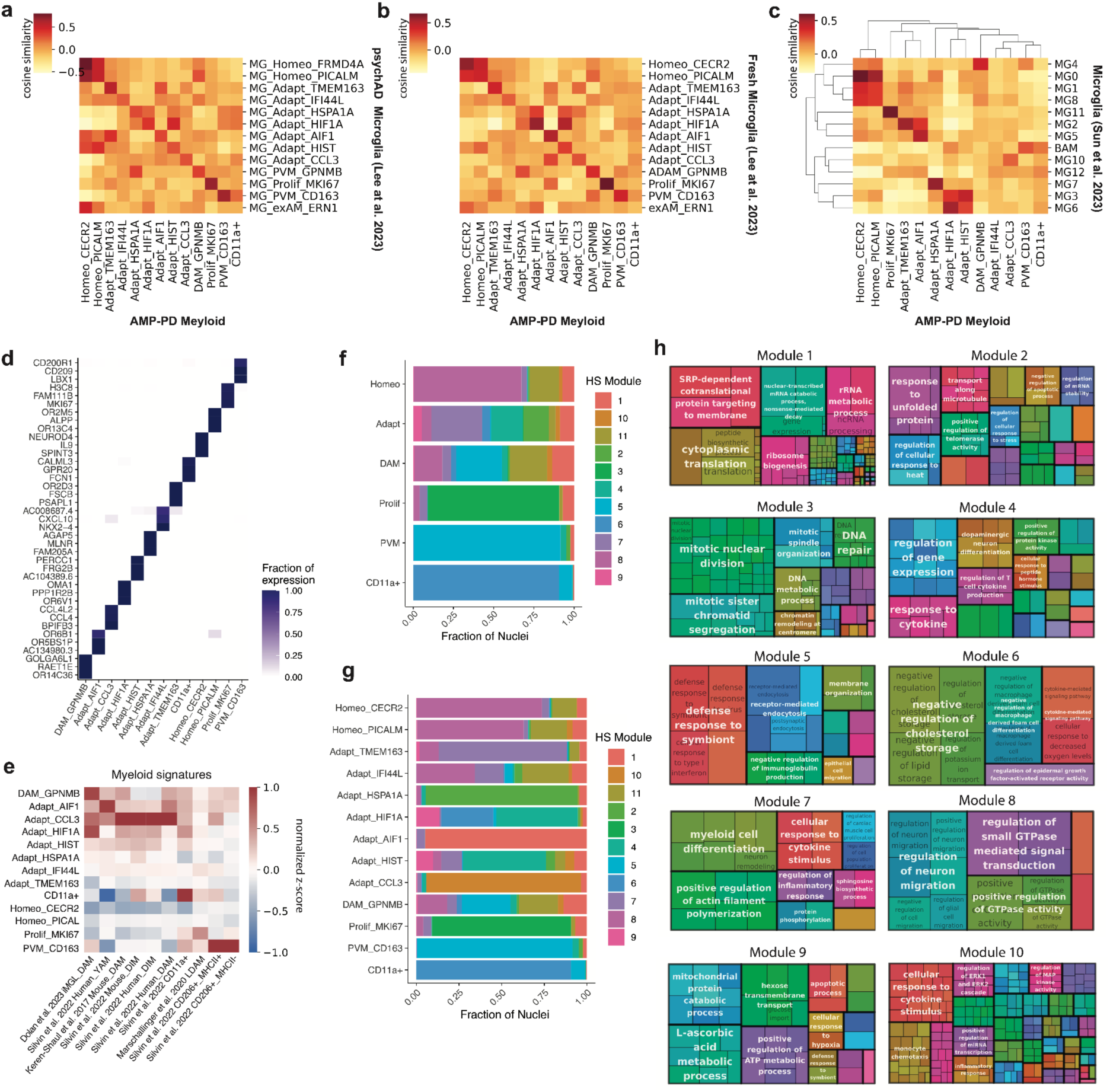
Functional Characterization and Cross-Dataset Mapping of Myeloid Subtypes in AMP-PD. (a-c) *Cosine similarity analysis comparing AMP-PD myeloid pseudobulk profiles to previously published datasets*: (a) cross-disorder microglial populations from Lee et al. (2024)^20^, (b) freshly isolated microglia from Lee et al. (2023)^67^, and (c) microglial clusters from Sun et al. (2023)^68^. (e) *Top three gene markers per AMP-PD myeloid subtype*, selected based on cell-type specificity (see **Methods**). (f) *Heatmap showing z-score–normalized enrichment of curated microglial gene signatures*^17,66,70,71^, across AMP-PD myeloid subtypes. **(f-g)** *Stacked bar plots showing the proportion of nuclei assigned to each hotspot* module across myeloid subclasses (f) and subtypes (g). **(h**) *Treemap summarizing biological themes* derived from GO Biological Process enrichment of hotspot modules (**f-g**), clustered by semantic similarity using the rrvgo package^31^.

**Extended Data Figure 7.**
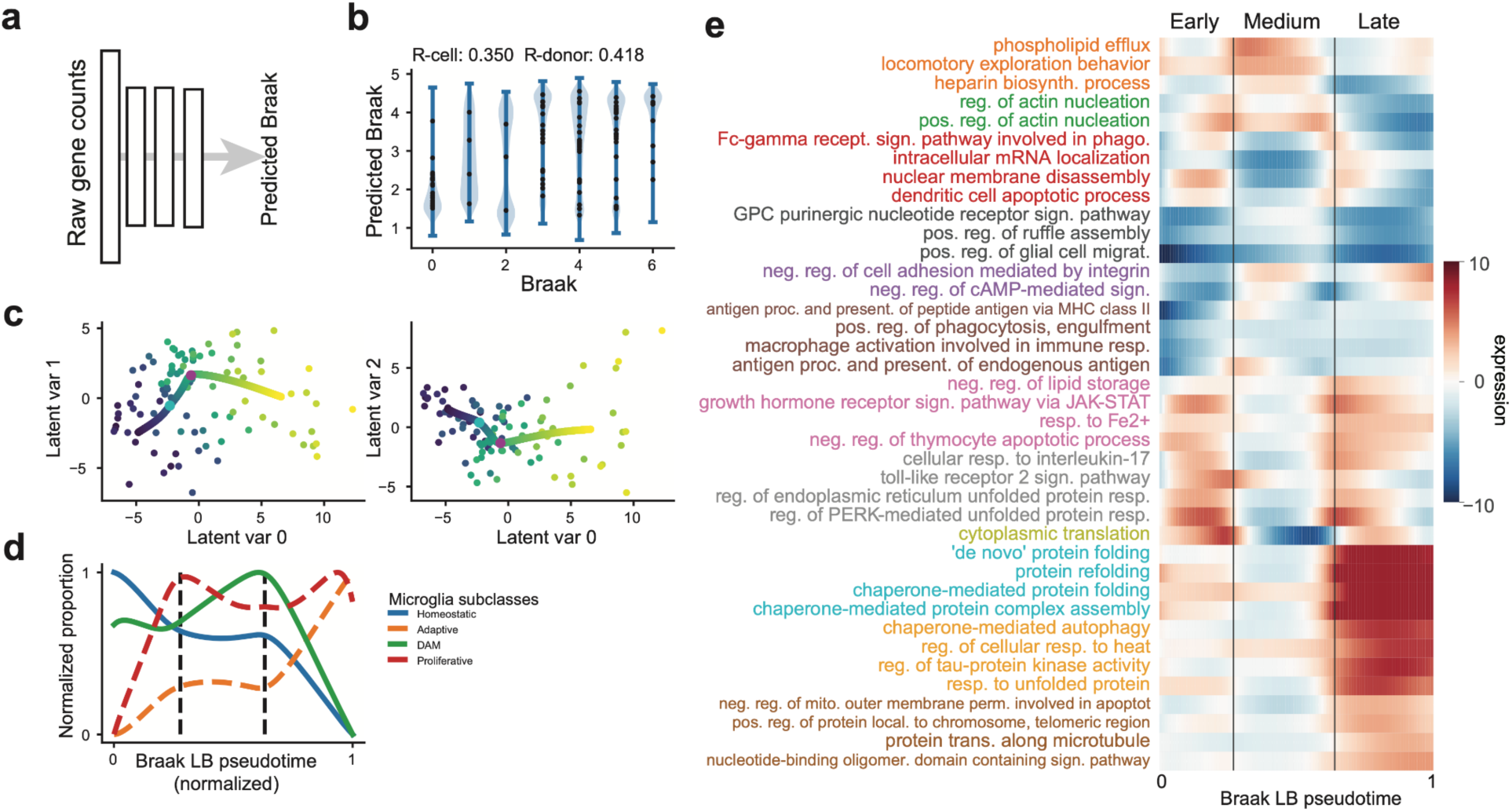
Modeling Braak LB Progression Using Gene Expression-Derived Pseudotime and Functional Signatures in Myeloid Cells. (a) *Schematic overview* illustrating the use of raw gene counts as input to train a neural network model of Braak LB stage using myeloid nuclei. **(b)** *Predicted Braak LB scores across known Braak LB stages*. The blue silhouettes show the distribution of cell level predicted Braak LB scores, and the black dots show the donor-averaged predicted Braak LB scores. The model achieved a moderate correlation with actual Braak LB staging (Pearson correlation at the cell and donor levels, respectively *r* = 0.35 and 0.41). **(c)** *Donor-averaged gene expression projected onto a low-dimensional latent space using PLS regression*. Each dot represents a donor, and the hue indicates its averaged predicted Braak LB score. The smoothed curve represents the gene trajectories projected onto the same latent space. The cyan and magenta circles indicate the nonlinear transition points (see **Methods**). **(d)** *Normalized proportions of microglial subclasses along the predicted Braak LB pseudotime*, highlighting subclass-specific progression patterns. The vertical dashed lines indicate the nonlinear transition points. **(e)** *Change in pathway gene expression calculated using a sliding window across predicted Braak LB pseudotime*. A curated list of pathways that met significance criteria (see **Methods**) were included. A full list of pathways that met the significance criteria is shown in **Supplementary** Fig. 12a. Hue indicates z-score values. Vertical lines are the transition points corresponding to Early, Medium and Late Braak LB stage, respectively. Font color indicates pathway cluster membership, in which pathways with similar transcriptional dynamics were grouped together (see **Methods**).

